# Characterizing individuals fulfilling clinical criteria for LATE in a tertiary memory clinic

**DOI:** 10.1101/2025.10.22.25338577

**Authors:** Colin Groot, Ismael L. Calandri, Ilse Bader, Diana I. Bocancea, Hannah de Bruin, Maria Carrigan, Lyduine E. Collij, Floor H. Duits, Suzie Kamps, Lotte A. de Koning, Afina W. Lemstra, Sophie E. Mastenbroek, Roos M. Rikken, Bastiaan G. J. van Tol, Marie R. Vermeiren, Alex Wesseling, Ye Xia, Charlotte E. Teunissen, Elsmarieke vd Giessen, Frederik Barkhof, Laura Jonkman, Casper de Boer, Annemieke Rozemuller, Anne E Leeuwis, Annelies E van der Vlies, Betty M. Tijms, Wiesje M. van der Flier, Yolande A. L. Pijnenburg, Emma M. Coomans, Rik Ossenkoppele

**Author notes:** Corresponding authors: Colin Groot, PhD, Alzheimer center Amsterdam, Amsterdam UMC, location VUmc, de Boelelaan 1117, 1081 HV, Amsterdam, the Netherlands Rik Ossenkoppele, Alzheimer center Amsterdam, Amsterdam UMC, location VUmc, de Boelelaan 1117, 1081 HV, Amsterdam, the Netherlands. Equally contributed as senior authors (Emma M. Coomans, Rik Ossenkoppele).

## Abstract

Limbic-predominant age-related TDP-43 encephalopathy (LATE) is characterized by an amnestic-and limbic-predominant phenotype, which can mimic the clinical presentation of Alzheimer’s disease (AD). Differentiating between LATE and AD is imperative in the era of disease-modifying treatments for AD, but there are no established biomarkers to detect TDP-43. In this observational study, we characterized individuals fulfilling the recently proposed clinical criteria for LATE in a tertiary memory clinic setting in terms of cognition, atrophy, and disease course.

From the Amsterdam Dementia Cohort (ADC), we included 3606 individuals aged above 50 years old who had a diagnosis of MCI or dementia. We first classified individuals based on criteria for memory impairment, atrophy, amyloid-status and tau-status, into (Probable and Possible) LATE, co-occurring LATE and AD (Possible LATE-AD), and AD without LATE. Next, we compared these four groups on demographic, clinical, cognitive, and brain atrophy data. Longitudinal cognitive data were available for a subset.

We classified 56/3606 (1.6% of the total group) as Probable LATE, 115 (3.2%) as Possible LATE, 127 (3.5%) as Possible LATE-AD, and 1675 (46.5%) as AD. The remaining 1633 (45.3%) were not classified within this framework (e.g., frontotemporal dementia). At baseline, individuals with Probable LATE had lower memory scores but higher scores on all other domains and MMSE than AD, while individuals with Possible LATE-AD had worse scores across all cognitive scores than AD (p<0.05). Individuals with Probable LATE progressed slower than AD on MMSE (sβ[SE]=0.12[0.05], p=0.02), memory (sβ[SE]=0.24[0.1], p=0.01), attention (sβ[SE]=0.34[0.17], p=0.05), and executive functioning (sβ[SE]=0.19[0.09], p=0.03). Individuals with Possible LATE-AD progressed faster than AD on MMSE (sβ[SE]=-0.12[0.05], p=0.01), attention (sβ[SE]=-0.35[0.17], p=0.04), and executive functioning (sβ[SE]=-0.2[0.09], p=0.03). Mortality risk, compared to AD, was lower in individuals with Probable LATE (HR=0.70 [0.49-0.99], p=0.04) and higher in Possible LATE-AD (HR=1.25 [1.01-1.53], p=0.04). Compared to AD, at baseline, individuals with Probable and Possible LATE had higher inferior temporal-to-hippocampal ratios (indicating limbic-predominant atrophy; β[SE]=0.27[0.08], p<0.01; β[SE]=0.38[0.11], p<0.01), and Probable LATE additionally showed smaller amygdalar volumes (β[SE]=-2.36[0.75], p<0.01). Individuals with Possible LATE-AD had thinner cortex at baseline in an “AD-signature” composite region compared to AD (β[SE]=-2.14[0.64], p=0.01; β[SE]=-3.26[0.96], p<0.01).

Among individuals from a tertiary memory clinic (mean age: 66 [6]), 8.2% were classified as Possible LATE, Probable LATE, or Possible LATE-AD. Probable and Possible LATE were characterized by a milder disease course than AD, while LATE-AD was characterized by a more aggressive disease course. These differential clinical trajectories highlight the prognostic utility of clinically defining LATE *in vivo*.

## Introduction

Limbic predominant age-related TDP-43 encephalopathy (LATE) is a pathologic entity that is associated with the pathologic finding of TAR DNA-binding protein 43 (TDP-43) proteinopathy in limbic brain regions^1^, a finding that is termed LATE neuropathological change (LATE-NC). The specific pathologic pattern in LATE-NC is distinct from TDP-43 subtypes of fronto-temporal lobar degeneration (FTLD)^2^ amyotrophic lateral sclerosis (ALS) and motor neuron disease (MND)^3^. During life, individuals with LATE-NC typically display an amnestic profile that mimics the early stages of Alzheimer’s disease (AD), which complicates clinical differentiation between LATE and AD^4^. This is reinforced by the radiological features typical of LATE, i.e., limbic-predominant atrophy, which is also characteristic of (early) AD^5, 6^. This neurobiological fingerprint closely expresses the localization of pathology in both LATE (TDP-43 proteinopathy) and AD (neurofibrillary tau tangles), which both have a propensity for limbic brain regions^1, 7^. While intracellular TDP-43 inclusions and their harmful effects on the brain have been recognized for some years^8^, TDP-43 proteinopathy was, at first, more commonly associated with FTLD and ALS. More recently, the high prevalence of LATE-NC and its clinical consequences became apparent based on assessments in clinico-pathologic cohorts^9–11^. Estimates indicate that LATE-NC might be present in up to 30% of individuals over 85^11, 12^, and may even exceed the prevalence of ADNC in individuals over 90^4, 12, 13^.

LATE-NC often co-occurs with AD pathology, especially with advancing age^1, 11, 14–16^. The co-occurrence of LATE-NC and AD pathology acts synergistically and results in worse clinical outcomes than isolated AD, while LATE-NC without AD pathology seems to result in slower clinical progression than “pure” AD or “pure” LATE^1, 10, 11, 15, 17, 18^. While clinically similar, differences between LATE and AD have been reported. For example, LATE is characterized by a sustained amnestic-predominant profile until later stages of the disease, while individuals with AD generally progress faster to multi-domain impairment^10^. To date, there are no established biomarkers with molecular specificity for TDP-43 pathology.

Therefore, expert consensus criteria were recently proposed that provide guidelines for clinically diagnosing LATE^14^. These criteria are based on the key characteristics of LATE, i.e., an amnestic clinical profile and a limbic-predominant neurodegeneration pattern, in conjunction with AD (i.e., amyloid-β and tau) biomarkers^14^. Here, we retrospectively applied the proposed clinical consensus criteria for LATE to patients with MCI or dementia from a tertiary memory clinic cohort, closely resembling the intended real-world clinical use of these criteria. Using neuropsychological, MRI, and biomarker data, we classified individuals into groups of (Probable and Possible) LATE, co-occurring LATE and AD (i.e., Possible LATE-AD), and AD (without LATE). We then assessed their demographic characteristics (age, sex, education, *APOE*ε4 genotype), clinical characteristics (cross-sectional and longitudinal cognition, mortality rates), and MRI (cross-sectional and longitudinal brain atrophy).

## Materials and Methods

### Study Participants and Procedures

We used data collected between September 1997 and March 2024 from the Amsterdam Dementia Cohort (ADC), consisting of individuals who visited the memory clinic of the Alzheimer Center Amsterdam and gave permission to use their data for research purposes. Detailed study procedures have been described previously^19, 20^. All patients underwent a diagnostic workup that included an assessment of medical history, a neurological exam, neuropsychological testing, brain MR, lumbar puncture, and venipuncture to allow for *APOE* genotyping. Positivity for amyloid-β was determined by amyloid-β-PET or CSF. Amyloid-β-PET was performed using [^11^C]Pittsburgh compound-B, [^18^F]florbetaben, [^18^F]florbetapir, or [^18^F]flutemetamol, and classified as negative (A-) or positive (A+) based on visual read methods for each tracer. In CSF, evidence for the presence of amyloid-β pathology (i.e., amyloid-positivity; A+) was based on a cut-off value of <813 pg/mL (Innotest, until June 2018)^21^ or <1,092 pg/mL (Elecsys, from June 2018 onward)^22^. Amyloid-data was available for 2856/3606 individuals (79.2%; 775 negative and 2081 positive). Tau positivity (T+) was determined in CSF and was based on hyperphosphorylated tau (ptau)181 concentrations of >19 pg/mL (Elecsys) or, in a limited number of cases (n=52), [^18^F]flortaucipir-PET visual read. Tau-data was available for 2715/3606 individuals (75.3%; 872 negative and 1843 positive). For the present study, we only selected individuals who were >50 years of age with cognitive impairment (i.e., a syndrome diagnosis of mild cognitive impairment [MCI] or dementia [see Table 1]^23–30^), but individuals with a non-neurodegenerative condition (e.g. delirium or brain tumor) or a primary psychiatric disorder (e.g., schizophrenia or major depressive disorder) were not included. This resulted in an initial screening sample of 4042 individuals. To be eligible for classification into our target groups (see “Operationalization of LATE criteria and Group Classification” and Fig. 1), baseline neuropsychological test data needed to be sufficient to generate a composite score for memory and for two other cognitive domains (i.e., attention, executive functioning, language and/or visuospatial ability), to allow for determination of amnestic or multi-domain impairment (see “Cognition” section). Of the 4,042 individuals, 88 (2.2%) did not meet this criterion. Furthermore, to determine a limbic-predominant radiological phenotype, individuals needed to have a baseline MRI scan that was of sufficient quality for visual assessment of medial temporal atrophy (MTA^6, 31^), along with an assessment of parietal atrophy (PA) and/or global cortical atrophy (GCA^32^; see “Neuroimaging processing” section). Of the remaining 3954 individuals, 348 (8.8%) did not meet this criterion, and, therefore, 3,606 individuals were eligible for potential LATE classification.

**Figure 1.**
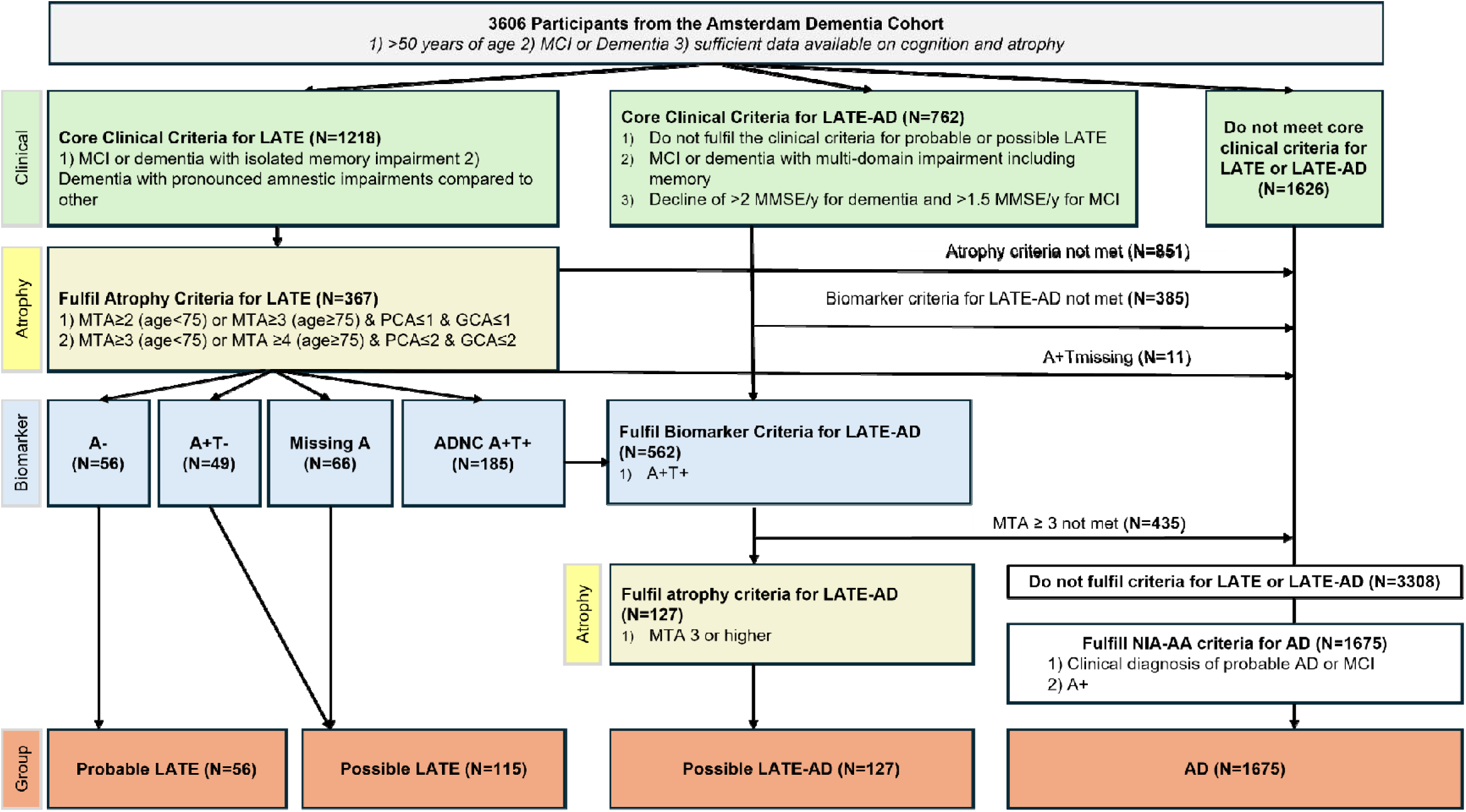
Flowchart of participant classification steps LATE=limbic-predominant age-related TDP-43 encephalopathy; AD=Alzheimer’s disease; MTA=medial temporal atrophy; PA=posterior atrophy; GCA=global cortical atrophy; A=amyloid-β biomarkers; T=hyperphosphorylated tau biomarkers

**Table 1:** Demographics across groups. Values are *n* (%) for categorical variables and mean (standard deviation) for continuous variables. P-values for overall group differences obtained using independent samples t-tests, Fisher’s exact tests and Kruskal Wallis tests (applied where appropriate). Pairwise differences between all groups are displayed in Supplementary Table 3. *-based on multidisciplinary consensus, #= measured on the qualitative Verhage scale from 1-7. Missing data; education *n* = 16; APOEε4 status *n* = 146; MMSE *n* = 35; MMSE yearly decline *n* = 141; NPI = 326. AD = Alzheimer’s disease; APOE = apolipoprotein E; DLB = Lewy body dementia; FTD = frontotemporal dementia; IQR=interquartile range; LATE = limbic-predominant age-related TDP-43 encephalopathy; MCI = mild cognitive impairment; MMSE = mini-mental state examination; VaD = vascular dementia.

|  | **AD** | **Probable LATE** | **Possible LATE** | **LATE-AD** | **p** |
| --- | --- | --- | --- | --- | --- |
| **N** | 1675 | 56 | 115 | 127 |  |
| **Age, years** | 66.07 (7.08) | 66.00 (6.38) | 69.86 (6.23) | 70.14 (6.77) | <0.001 |
| **Female, n(%)** | 840 (50.1) | 38 (67.9) | 55 (47.8) | 70 (55.1) | 0.043 |
| **Education#** | 5.12 (1.28) | 4.69 (1.17) | 5.09 (1.24) | 5.09 (1.34) | 0.106 |
| **MMSE** | 22.58 (4.94) | 24.13 (3.10) | 21.70 (4.11) | 19.00 (5.55) | <0.001 |
| **NPI total** | 11.56 (12.25) | 11.80 (11.23) | 14.53 (15.56) | 13.11 (13.54) | 0.130 |
| **Suspected etiology\*** |  |  |  |  | <0.001 |
| AD | 1217 (72.7) | 20 (35.7) | 96 (83.5) | 115 (90.6) |  |
| FTD | 0 (0.0) | 15 (26.8) | 4 (3.5) | 6 (4.7) |  |
| VaD | 0 (0.0) | 0 (0.0) | 5 (4.3) | 0 (0.0) |  |
| DLB | 0 (0.0) | 2 (3.6) | 1 (0.9) | 0 (0.0) |  |
| MCI | 458 (27.3) | 16 (28.6) | 8 (7.0) | 6 (4.7) |  |
| Other | 0 (0.0) | 3 (5.4) | 1 (0.9) | 0 (0.0) |  |
| **APOEε4, n(%) carrier** | 1100 (70.0) | 20 (40.0) | 58 (64.5) | 82 (70.7) | <0.001 |

### Cognition

Included individuals underwent a comprehensive neuropsychological test battery, and test scores were categorized into 5 cognitive domains (memory, attention, language, executive functioning [EF], and visuospatial ability) by expert neuropsychologists at the Alzheimer center Amsterdam^33^. Memory was assessed with the Dutch version of the Rey Auditory Verbal Learning Test (RAVLT), the immediate and delayed recall conditions, as well as part A (two trials) of the Visual Association Test (VAT). We assessed attention with the following tests: Trail Making Test (TMT)-A, digit span forward, Stroop card 1, and card 2. Language was measured with the naming condition of the VAT and the animal fluency (60-second test). Note that we did not include the animal fluency test when determining the diagnostic groups, as it also relies on semantic memory, and the clinical criteria for LATE^14^ outline that LATE cases may have semantic memory impairment based on autopsy study data that revealed semantic memory deficits in individuals found to have LATE-NC^10, 34^ (see Supplemental Table 1-2 for further details). Hence, the language domain scores that included both the naming condition of the VAT and animal fluency were only used in the longitudinal analyses to determine the decline in this domain, but not in the LATE classification. We assessed EF with the letter fluency test, Stroop card 3, digit span backward, and the Frontal Assessment Battery (FAB). Visuospatial ability was assessed with the following tests from the Visual Object and Space Perception battery (VOSP): fragmented letters, dot counting, and number location. Scores that were based on the time taken to complete the test (i.e., Stroop and TMT) were inverted so that lower scores indicated worse performance. To account for missing data, we imputed all neuropsychological data with the *mice* package from R using the entire dataset of neuropsychological tests along with MMSE, age, sex, education, and clinical diagnosis as predictors. To allow the calculation of composite cognitive domain scores, all raw test scores were then converted into *z* scores based on the mean and SD from independent normative data from 440 cognitively unimpaired (CU) amyloid-β-negative participants from the ADC (mean age 59.9 years [*SD* = 6.05], 40% female)^35^. The z-scores of tests within each domain were then averaged to generate the composite scores. Cognitive domain scores were only computed if an individual had at least 1 test within that domain available in the non-imputed dataset to avoid composite scores being entirely composed of imputed scores. The metric of the resulting cognitive domain composite scores represents the SD from the independent control group (e.g.,-1 indicates performance of 1 SD below normal functioning), and we determined ≤-2 to signify a significant domain impairment.

As a measure of global cognitive functioning, we implemented MMSE scores. To provide an estimate of global cognitive decline *prior* to diagnosis, we additionally computed an MMSE yearly decline metric that is obtained by subtracting the MMSE contemporaneous to diagnosis from 30 (max score for the MMSE) and then dividing by the self-reported symptom duration.

### Operationalization of LATE criteria and Group Classification

The consensus criteria outlined by Wolk et al., 2024^14^, classify individuals into 3 separate LATE groups in a probabilistic manner based on whether they are suspected to have LATE-NC using clinical, radiological, and biomarker profiles. Probable LATE has a clinical-radiological profile fulfilling LATE criteria with negative amyloid-biomarkers. This group is deemed most likely to have LATE-NC as the driving etiology (i.e., “pure LATE”). Possible LATE has a clinical-radiological profile fulfilling LATE criteria with positive or missing amyloid biomarkers. Possible LATE-AD has a clinical-radiological profile fulfilling LATE criteria in addition to positive amyloid-and tau-biomarkers (i.e., suspected LATE-NC/ADNC co-pathology). The specific required and supporting criteria for every one of these categories are outlined in Supplemental Tables 1 and 2, and in the original publication^14^. In this study, the consensus criteria outlined by Wolk et al. (2024)^14^ were adapted to accommodate the data available in the Amsterdam Dementia cohort. The specific operationalization of each of the criteria is also detailed in Supplemental Tables 1 and 2, and in Fig. 1. Briefly, an individual was classified as LATE when there was an isolated memory impairment, or, for dementia cases only, a cognitive profile that was characterized by a disproportionate memory impairment relative to the other cognitive domain scores of that individual, i.e., memory is more than 1SD below the average across all domains. The latter accounts for a possible amnestic-predominant but multi-domain impairment in later LATE disease stages. A radiological phenotype fitting with LATE was defined by age-adjusted visual read measures of atrophy determined on structural MRI; medial temporal atrophy (MTA), posterior atrophy (PA), and global cortical atrophy (GCA)^6, 32^. Consistent with criteria for AD, the radiological criteria for LATE do not define a strict cut-off when it comes to classifying a limbic-predominant atrophy profile. In our operationalization, individuals could have 1) MTA≥2 (when aged <75) or MTA≥3 with PA and/or GCA ≤1, or 2) MTA ≥3 (aged >75) or MTA ≥4 with GCA and/or PA 2. As outlined in the Wolk et al publication^14^, among individuals who adhered to these clinical and radiological profiles, those with negative amyloid-biomarkers (i.e., A-) were categorized as “Probable LATE”. Individuals with missing A biomarkers (A_missing_) or with an A+T-biomarker profile were categorized as “Possible LATE”. The individuals who adhered to the clinical and radiological profiles for LATE but who were A+T+ (and therefore not eligible to be classified as Probable or Possible LATE) were pooled with A+T+ individuals with a multi-domain impairment (including memory) for further classification. Among this combined group, those with MTA ≥3 were classified as “Possible LATE-AD”, henceforth referred to as “LATE-AD” to avoid confusion with the Possible LATE group. A+T+ individuals who did not meet the MTA ≥3 criterion for LATE-AD, were grouped with all A+ individuals who did not meet clinical and radiological profiles for any of the LATE groups and were classified into the AD reference group if they also received a clinical diagnosis of AD based on a multidisciplinary consensus meeting. The flow chart in Fig. 1 describes the participant classification steps in greater detail.

### Mortality

Survival was defined as the time from the screening visit at the Alzheimer Center Amsterdam to the event (being either death or censoring). Data on mortality was collected from the Central Public Administration until September 2024 and included the status (dead/alive) and date of death. If data from the Central Public Administration were not available, the date of the last visit to the Alzheimer Center Amsterdam was included as the censoring date.

### Neuroimaging processing

We acquired structural T1-weighted MR images according to standardized acquisition protocols described previously^36^. Regional cortical thickness and surface area from regions of the Desikan-Killiany atlas were obtained in subject-space using *FreeSurfer* (version:7.1; http://freesurfer.net/). We also derived subcortical and cortical volumetric measures using automatic segmentation (ASEG), distributed as part of *FreeSurfer*. Since data were acquired across multiple scanners and field strengths (i.e., 1T, 1.5T, and 3T), we applied the harmonization procedure *neurocombat,* to facilitate the pooling of data across scanners^37^.

*Neurocombat* accounts for variance in the MRI data caused by differences in scanner field strength while preserving biological variation accounted for by patient characteristics (i.e., age and sex). Tailored *FreeSurfer*-based longitudinal image analysis pipelines were used to process MRI scans used for longitudinal analyses. For image series within individuals, we only selected scans with the same field strength to avoid spurious longitudinal findings resulting from differences in field strength.

In order to cover regions typically associated with LATE or AD (or both) we selected the following regions-of-interest (ROI) to assess: hippocampal volume, amygdalar volume, entorhinal thickness, AD signature cortical thickness (weighted average of entorhinal, inferior temporal, middle temporal, parahippocampal, rostral middle frontal, medial orbitofrontal, precuneus, inferior parietal, and supramarginal cortical thickness), and a ratio between inferior temporal thickness / hippocampal volume (ITH ratio; smaller values indicate relative limbic vs neocortical atrophy), which has been proposed to distinguish LATE from AD^38–40^. We will present results for the AD signature ROI, amygdalar ROI and the ITH ratio in the main text, and the rest of the ROIs are displayed in the supplemental material.

## Statistical analyses

All statistical analyses and visualizations were performed in *R* software (version:4.3.3).

We fitted linear regression models to examine cross-sectional differences between groups on cognition and atrophy, and we fitted linear mixed models (LMMs) to examine differences in longitudinal trajectories in cognitive domains scores and brain atrophy. Models were adjusted for age, sex, and education (for models with cognition as the outcome). LMM models also included a group*time interaction effect. For LMM, we fitted models with and without random slopes and intercepts and selected the best-fitting model based on the lowest Akaike Information Criterion (AIC) value. For the survival analyses, raw median survival estimates were first obtained from Kaplan-Meier analysis for survival from the time of dementia screening to the event (i.e., death or censoring). Cox proportional-hazard models were performed to evaluate whether differences in mortality risk were statistically significant when correcting for the potential confounders age, sex, education, and syndrome diagnosis.

Across all analyses, we will focus on differences between the LATE groups and AD in the main text, while pairwise differences between all groups are outlined in the supplement.

### Standard Protocol Approvals, Registrations, and Patient Consents

Written informed consent was obtained for participation in the ADC, and study procedures were approved by the institutional review board of the Amsterdam UMC (REC 2017.315). This study was performed in accordance with the World Medical Association Declaration of Helsinki, Ethical Principles for Medical Research Involving Human Subjects 2013. The handling of data was in agreement with the EU General Data Protection Regulation and the Dutch Act on Implementation of the General Data Protection Regulation.

## Data Availability Statement

Anonymized data supporting the findings of this study are accessible to a qualified investigator upon reasonable request.

## Results

Following the participant classification steps outlined in Fig. 1, we classified 56/3606 (1.6% of all eligible individuals) as Probable LATE, 115 (3.2%) as Possible LATE, and 127 (3.5%) as LATE-AD, totaling 298 (8.2%) individuals being classified into one of the clinical LATE groups. Of the 3,308 participants who did not fulfill criteria for Probable LATE, Possible LATE, or LATE-AD, 1675 participants (46.5%) were amyloid-positive and had a clinical diagnosis of MCI or probable AD, and were categorized into our AD reference group. The remaining 1,633 (45.3%) individuals did not meet our criteria for either LATE or AD and were not included in the rest of our analyses. This group consisted of 380 individuals with a clinical diagnosis of AD (but missing or negative amyloid biomarkers), 457 MCI (but missing or negative amyloid biomarkers), 264 frontotemporal dementia, 132 vascular dementia, 234 dementia with Lewy bodies, or 166 with another (or unspecified) dementia diagnosis. Table 1 displays the demographic and clinical characteristics of the groups. The differences between the LATE groups and AD are described here, while all pairwise differences between groups are given in Supplementary Table 3. The Probable LATE group had a higher percentage of females (p<0.01) and a lower prevalence of APOEε4 compared to AD (p<0.01). The Possible LATE and LATE-AD groups were older than the AD group (all p<0.001), and the LATE-AD groups additionally had a lower baseline MMSE than AD (p<0.001).

### Cognition

Group differences described here will focus on differences between AD and the LATE groups, all pairwise differences between groups are provided in Supplementary Table 4. Since baseline cognitive scores were part of our group classification, differences in baseline cognitive scores between groups should be regarded as descriptive and are only outlined in detail in Fig. 2 and Supplementary Table 4. In brief, baseline memory was lower in all LATE groups compared to AD. Performance on the other cognitive domains was generally better for individuals with Probable LATE and worse for individuals with LATE-AD when compared to AD (Fig. 2).

**Figure 2.**
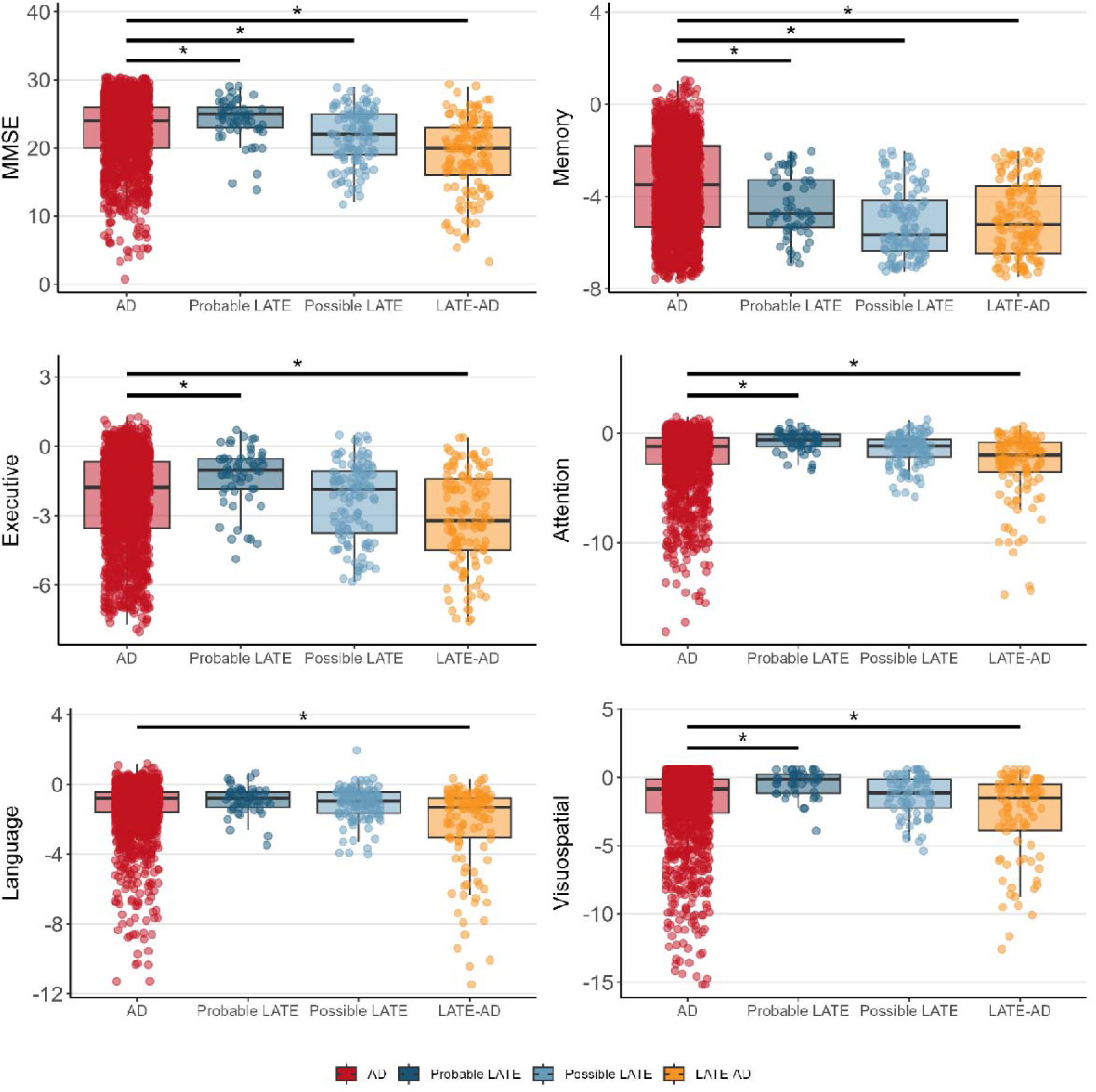
Baseline cognitive performance across diagnostic groups Differences between AD and the LATE groups in baseline cognition are indicated by the significance bars. All pairwise differences between groups are outlined in Supplementary Table 4.

For longitudinal decline, we observed that individuals with Probable LATE progressed slower than AD on MMSE (sβ[SE]=0.12[0.05], p=0.02), memory (sβ[SE]=0.24[0.1], p=0.01), attention (sβ[SE]=0.34[0.17], p=0.05), and executive functioning (sβ[SE]=0.19[0.09], p=0.03). Furthermore, individuals with LATE-AD progressed faster than AD on MMSE (sβ[SE]=-0.12[0.05], p=0.01), attention (sβ[SE]=-0.35[0.17], p=0.04), and executive functioning (sβ[SE]=-0.2[0.09], p=0.03). There were no differences in longitudinal cognitive performance between individuals with Possible LATE and AD (Fig. 3).

**Figure 3.**
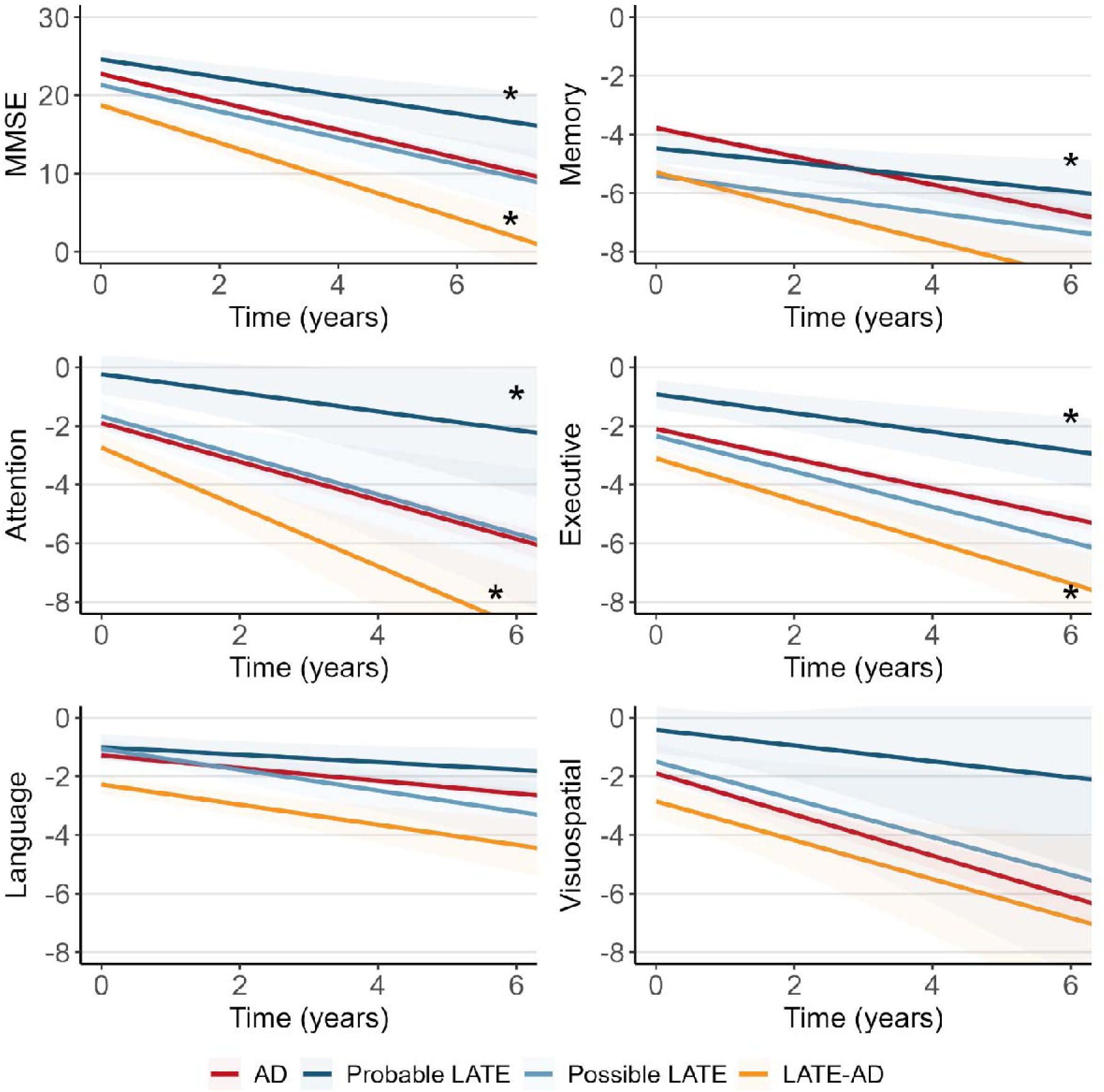
Longitudinal cognitive performance across diagnostic groups Differences between AD and the LATE groups in longitudinal cognitive performance (indicated by the slope) are indicated by an *. All pairwise differences between groups are outlined in Supplementary Table 4.

### Mortality

Out of the 1973 individuals included in the main analyses, 1187 (60.2%) died over the course of follow-up (mean follow-up time: 6.0[3.6]). The estimated median survival time was 9.0 years (95%CI: 8.3-12.2) for individuals with Probable LATE, 7.7 years (7.4-8.0) for AD, 5.8 years (4.7-7.3) for individuals with Possible LATE, and 5.7 years (5.0-6.6) for individuals with LATE-AD (Fig. 4). Cox proportional-hazards models showed that mortality risk, compared to AD, was lower in individuals with Probable LATE (HR=0.70 [0.49-0.99], p=0.04), whereas mortality risk was higher in the LATE-AD group (HR=1.25 [1.01-1.53], p=0.04). There was no difference in mortality risk between individuals with Possible LATE and AD (HR=0.85[0.68-1.07], p=0.174; Fig. 4). All pairwise differences between LATE groups are displayed in Supplementary Table 5.

**Figure 4.**
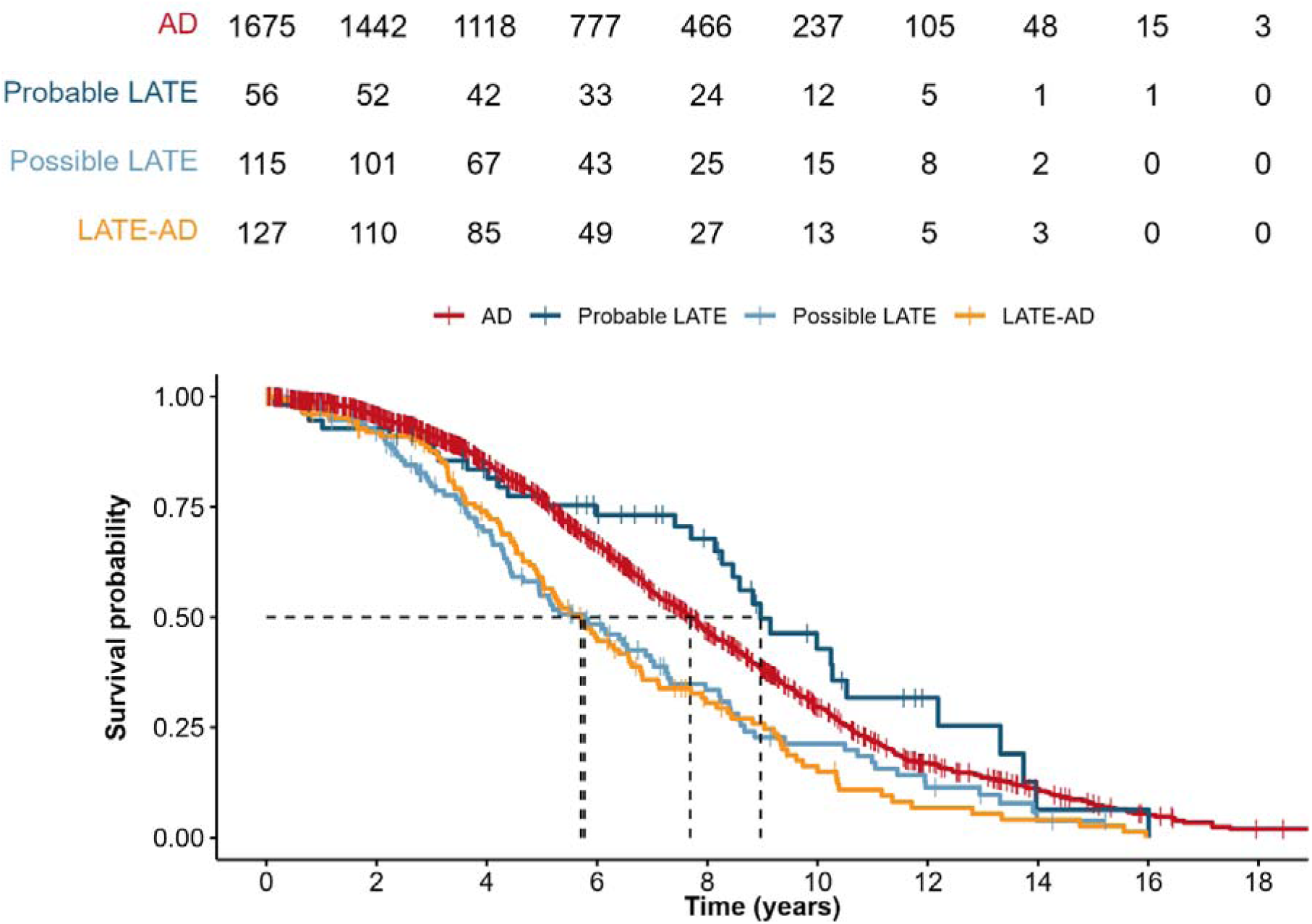
Mortality rates across diagnostic groups All pairwise differences between groups in HR are displayed in Supplementary Table 5. HR=Hazard ratio

### MRI

Group differences described here will focus on differences between AD and the LATE groups, all pairwise differences between groups are provided in Supplementary Table 6.

Individuals with LATE-AD had thinner cortex in an “AD-signature” composite region compared to AD at baseline (β[SE]=-3.26[0.96], p<0.01), indicating a greater degree of cortical atrophy. Individuals with Probable LATE showed smaller amygdalar volumes at baseline than AD (β[SE]=-2.36[0.75], p<0.01). Individuals with Probable LATE and Possible LATE had higher inferior temporal to hippocampal ratios (indicating limbic-predominant atrophy) compared to AD (β[SE]=0.27[0.08], p<0.01; β[SE]=0.38[0.11], p<0.01; Fig. 5).

**Figure 5.**
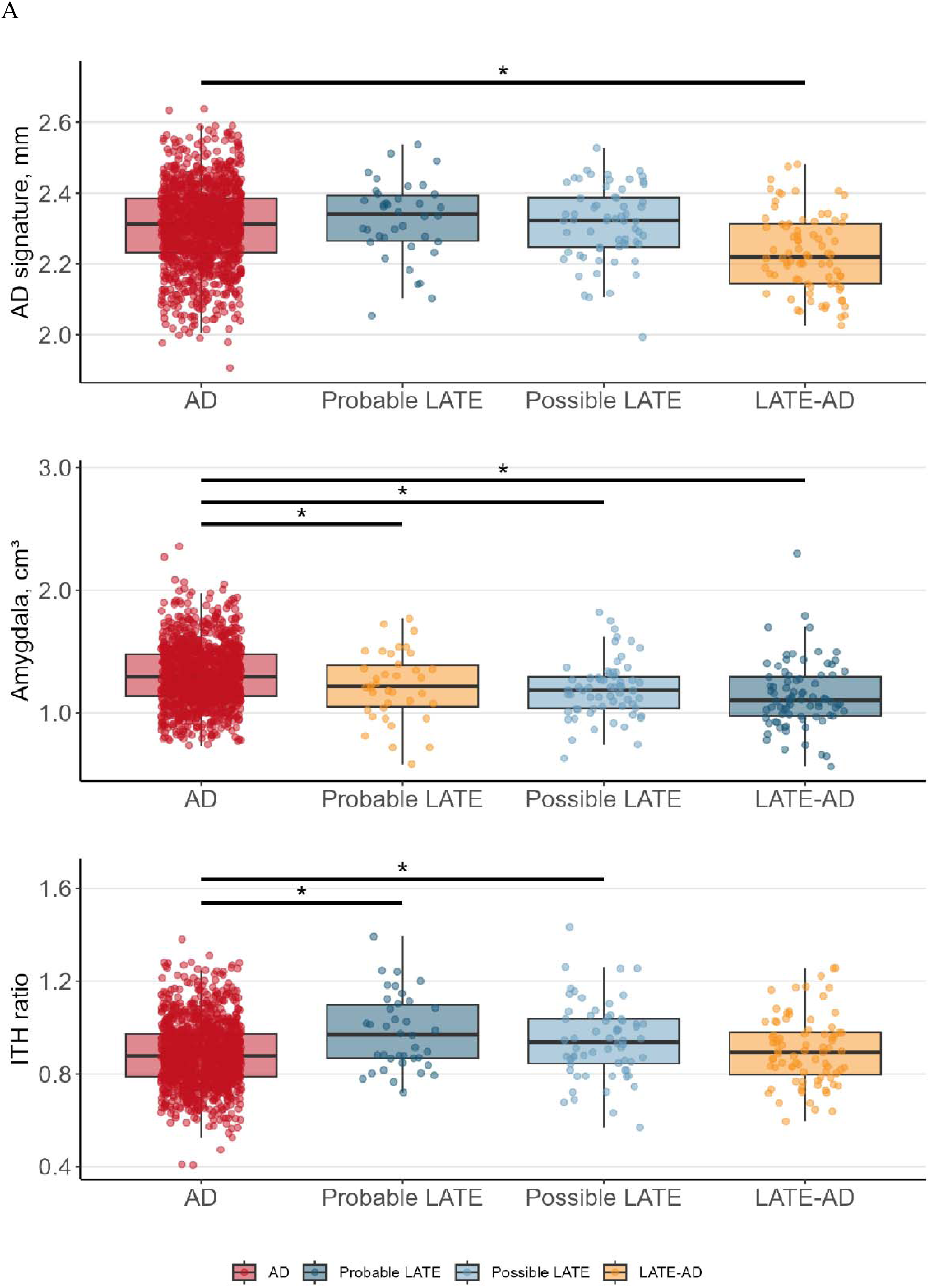
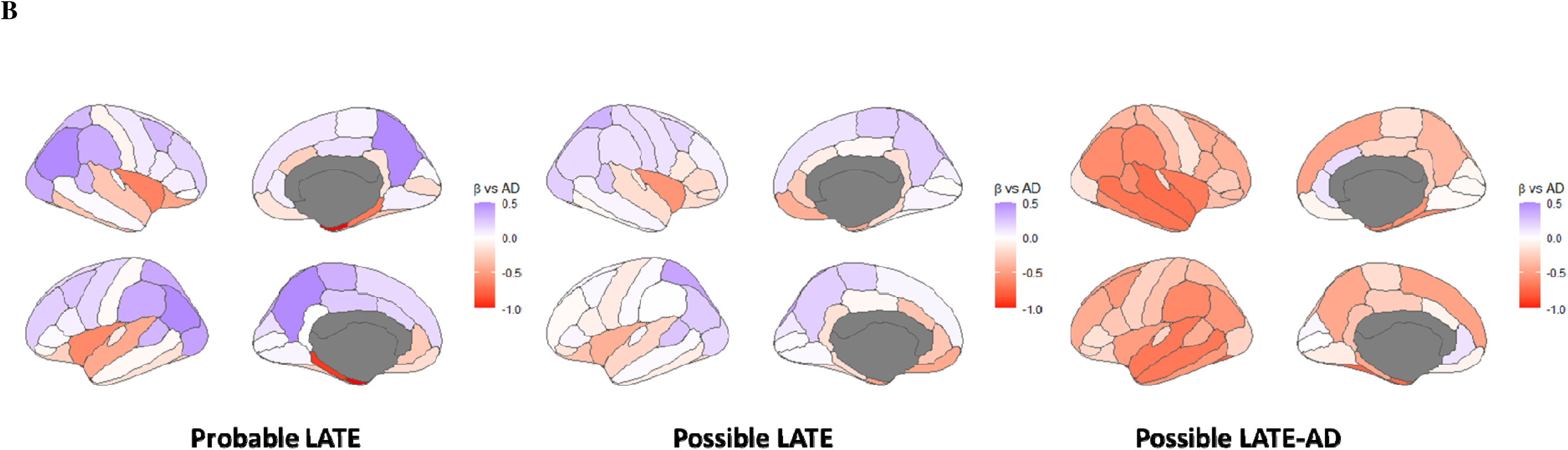
Baseline brain atrophy across diagnostic groups. (A) Bar graph showing group differences in baseline atrophy. Differences between AD and the LATE groups obtained using general linear models are indicated by the significance bars. (B) The surface projections are colored according to the β of the group effects on atrophy (i.e. LATE groups *vs* AD) from the general linear models assessing cross-sectional atrophy. All pairwise differences between groups are outlined in Supplementary Table 6 and results from additional regions of interest are displayed in Supplementary Fig. 1. ITH=inferior temporal thickness/hippocampal volume, higher values indicate more limbic-predominant atrophy

Compared to AD, individuals with Probable LATE, Possible LATE and LATE-AD had decreased hippocampal volumes at baseline (β[SE]=-4.30[1.26], p<0.01; β[SE]=-2.14[0.63], p=0.01; β[SE]=-2.33[0.65], p<0.01). Note that relative medial temporal atrophy was an inclusion criterion for the LATE groups, so these results were expected. Individuals with LATE-AD had thinner whole brain cortex compared to AD at baseline (β[SE]=-2.14[0.64], p=0.01; Supplementary Fig. 1).

We did not observe any significant differences in longitudinal atrophy rates between the groups (all p>0.05; Fig. 6).

**Figure 6.**
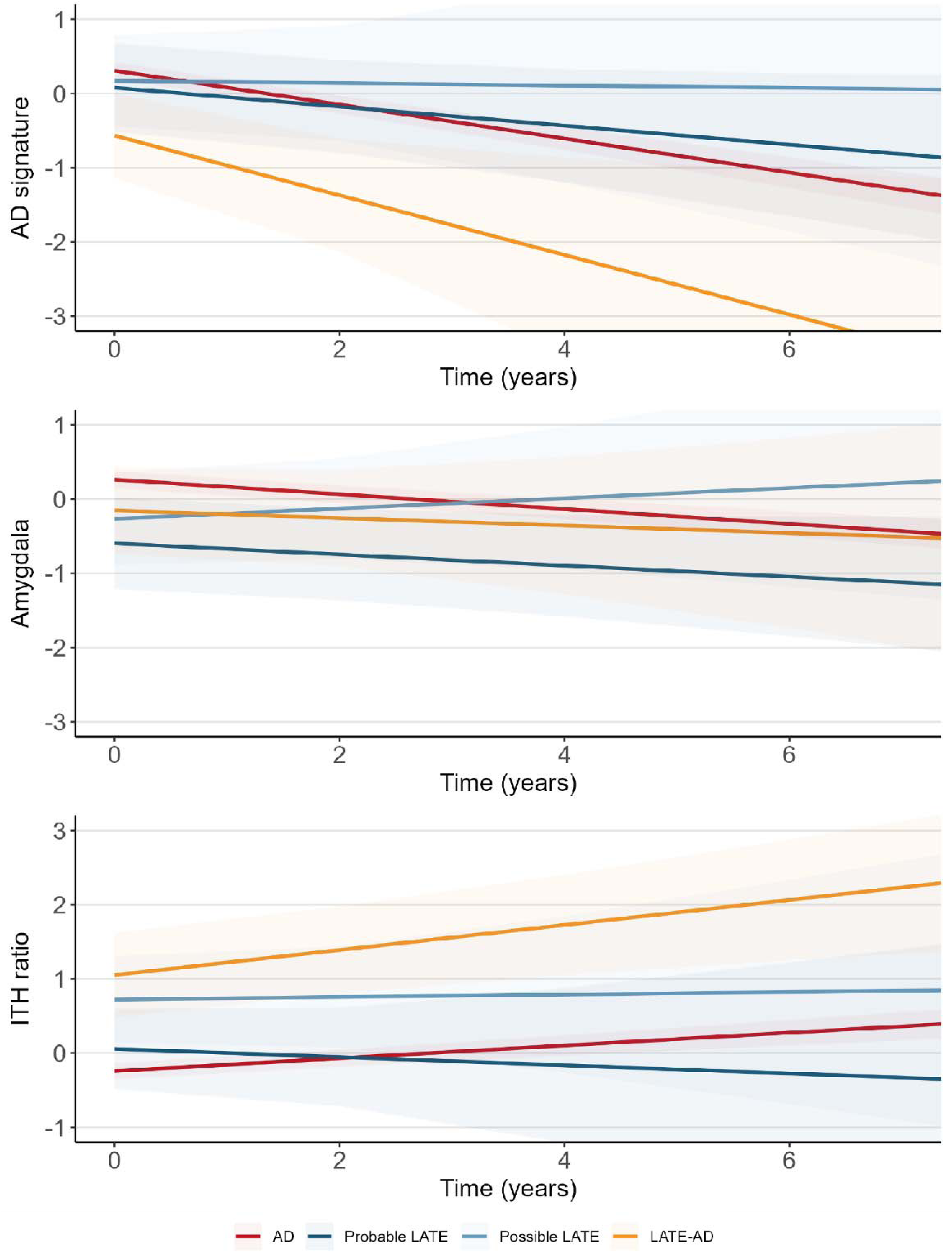
Longitudinal brain atrophy across diagnostic groups Line graphs indicate differences in longitudinal atrophy between groups. Differences between AD and the LATE groups obtained using linear mixed effects analyses (indicated by the slope) are highlighted with an *. Although the Possible and Probable LATE groups tended to have slower decline in AD-signature region cortical thickness (top graph), none of the time*group interaction effects were significant (p>0.05). All pairwise differences between groups are outlined in Supplementary Table 6, and results from additional regions of interest are displayed in Supplementary Fig. 1. ITH=inferior temporal thickness/hippocampal volume, higher values indicate more limbic-predominant atrophy.

## Sensitivity analyses

The Possible LATE group consisted of individuals with an A+T-biomarker profile (N=49), and those with missing A biomarkers (A_missing_; N=66). To assess possible differences between these two subgroups in our primary outcomes, we separated these two subgroups and assessed longitudinal cognition, mortality rates, and longitudinal atrophy. We found that individuals with Possible LATE A+T-had milder decline in language (β[SE]=-0.32(0.10), p<0.01; Supplementary Table 6) and faster atrophy rates in the hippocampus (β[SE]=0.67(0.31), p=0.03; Supplementary Table 8) than individuals with Possible LATE A_missing_. Similar to the separation of the Possible LATE group, we also separated the Probable LATE group into individuals with an A-T-biomarker profile (N=35) and those with an A-T+ profile (N=21).

Mortality risk was lower in A-T+ Probable LATE compared to the rest of the Probable LATE group (HR=0.35 [0.15-0.82], p=0.02). There were no differences in longitudinal cognition between these two Probable LATE subgroups (Supplementary Table 7), and atrophy could not be assessed because there were no A-T+ Probable LATE individuals with longitudinal MRI.

## Discussion

We applied the recently proposed consensus clinical criteria for LATE^14^ to individuals from a tertiary memory clinic. Based on their clinical, radiological and AD biomarker profile, 56 (1.6%) individuals could be classified as Probable LATE, 115 (3.2%) as Possible LATE and 127 (3.5%) as Possible LATE-AD, while 1675 participants (46.5%) adhered to clinical criteria for AD and were used as a reference group. We observed that individuals with Probable LATE generally showed slower cognitive decline than all other groups, and also showed lower mortality rates. The LATE-AD group, on the other hand, declined faster on all cognitive domains than AD and had higher mortality rates. Baseline inferior temporal cortical thickness to hippocampus volume (ITH) ratios differentiated individuals with Probable LATE and Possible LATE from AD, while amygdalar volumes were additionally smaller in individuals with Probable LATE compared to AD. Individuals with LATE-AD showed more severe cortical atrophy at baseline compared to AD. Taken together, among a tertiary memory clinic cohort, we were able to classify Possible LATE, Probable LATE, and LATE-AD groups that were not only distinct in their baseline clinical-radiological phenotypes but also showed unique clinical trajectories. These findings constitute the first assessment of the clinical utility of the newly proposed LATE criteria^14^ and highlight that the LATE classification can be highly relevant for diagnostic and prognostic purposes.

The primary challenge in differentiating LATE, especially from early AD, has been their overlapping clinical and radiological presentations, characterized by an amnestic-predominant and limbic-predominant profile^12–14, 16, 38^. The differentiation has been further complicated by a prevalent co-occurrence of LATE-NC and AD pathology, which may be related to shared risk factors or pathological synergy^1, 41^. In this study, we demonstrate that applying the newly proposed clinical criteria produced groups that were distinct from AD in clinical-radiological profiles as well as clinical trajectories. Among these, Probable LATE was characterized by the mildest clinical decline and lowest mortality rates, which is in line with previous literature^10, 13, 16, 42, 43^. While other pathologies may still be present to a degree in individuals classified as Probable LATE, LATE-NC is believed to be the dominant pathology^4, 34, 44^. In our own assessment, we were unable to confirm underlying TDP-43 pathology due to the limited availability of TDP-43 staining at post-mortem assessment.

However, given the negative amyloid-β biomarkers, AD pathology is unlikely to contribute significantly to symptoms in our Probable LATE group. This is further indicated by the relatively low prevalence of *APOE*ε4 (40%) in the Probable LATE group, compared to 70% in AD in our sample, which is an estimate for AD that is in line with previous studies^45^.

Although much more strongly associated with AD pathology, *APOE*ε4 has been reported to be a risk factor for LATE-NC, possibly relating to pathological synergy with AD^16, 46, 47^. The hypothesized association between *APOE*ε4 and LATE-NC might explain why *APOE*ε4 is still over-represented in our LATE groups compared to the general population (25% in Europe^45^). A clinical feature that has been consistently associated with LATE-NC is age, and age has even been incorporated into the limbic-predominant *age-related* TDP-43 encephalopahy (i.e., LATE) term. LATE has been hypothesized to predominantly affect individuals aged 75 and older^1, 48, 49^, whereas our cohort had a mean age of 66 (SD:6) years old. This may have contributed to the relatively low prevalence of LATE in our sample (8.2% for all LATE groups combined), whereas previous studies have reported higher percentages in older clinico-pathological cohorts (estimated to be 25%–40% in ≥85 years of age)^14^. Interestingly, however, we observed that individuals identified as Probable LATE (i.e. the group with the highest probability of LATE-NC as the primary etiology) in our memory clinic sample were not older than those in the AD group, and were indeed younger than the individuals with Possible LATE and LATE-AD groups (Supplementary Table 3). This suggests a more nuanced influence of age on the likelihood of developing a clinical presentation consistent with LATE, which should be examined in cohorts encompassing a sufficiently broad age range to allow direct assessment of age effects on the prevalence of clinically defined LATE. Another notable observation in the Probable LATE group was a higher proportion of male participants compared to AD. This male propensity for LATE was also observed in a previous study that proposed criteria for a related syndrome, limbic-predominant amnestic syndrome (LANS), where the male-to-female ratio was 7:1 in a subsample of individuals from ADNI who exhibited TDP-43 proteinopathy. However, this subsample was small (n=8), and the male overrepresentation was not observed in a Mayo Clinic cohort (n=5/9 males), which was also assessed in the same study^50^. While female sex was previously hypothesized to be related to LATE, given that females are more likely to reach advanced age and LATE is believed to be associated with older age^1^, our own, and previously reported, male over-representations suggests that further assessment is needed to explore sex *vs* LATE associations. In line with a previous investigation^51^, we observed that, at baseline, amygdalar and hippocampal atrophy in Probable LATE were more pronounced compared to AD, and the ITH ratio indicated more pronounced limbic-predominant atrophy in individuals with Probable LATE compared to AD. Given that medial temporal atrophy was used as a classification criterion in our operationalization of LATE, there is some degree of circularity in these results. However, it does highlight that these specific MRI measures may be of particular utility for classifying individuals with Probable LATE cases in the memory clinic.

In contrast to the Probable LATE group, the Possible LATE group showed similar clinical trajectories and mortality rates compared to AD, and *APOE*ε4 prevalence was similar. As also alluded to in the term *Possible* LATE, these results suggest that this group is not only neurobiologically (i.e., partly comprised of A+ individuals) but also clinically less distinct from “pure” AD. Our baseline atrophy measures revealed that the inferior-temporal cortical thickness to hippocampal volumes (ITH) ratios were different between individuals with Possible LATE and AD. Together with previous findings^39^, and an equivalent difference in the ITH ratio between individuals with Probable LATE and AD, this indicates that the ITH ratio metric could be a sensitive marker of LATE-NC, sensitive enough to distinguish Possible LATE from AD. Of note, the Possible LATE group was comprised of both individuals with amyloid-β-positive but tau-negative biomarkers (A+T-; possibly indicating incipient AD) and individuals with missing amyloid-β biomarkers (A_missing_) biomarkers (possibly indicating undetected AD). We show that the A+T-and A_missing_ subgroups of Possible LATE show different clinical profiles and trajectories. This highlights the importance of incorporating biomarkers into clinical criteria for LATE, and suggests that individuals with missing biomarkers may not fit well into the same category as individuals with a known biomarker profile. A further refinement of biomarker categories might also be pertinent to the Probable LATE group, where biomarkers were implemented to rule out AD pathology. In the current clinical criteria, this was based only on amyloid-β-biomarkers, while an A-T+ profile was eligible for classification into the Probable LATE group^14^. We showed that the A-T+ subgroup of individuals with Probable LATE did not have significantly different clinical-radiological profiles or longitudinal trajectories compared to the rest of the Probable LATE group. However, this might have been the result of a small sample size for the A-T+ Probable LATE subsample (N=21). The effects of primary age-related tauopathy (PART; defined by the A-T+ profile)^52, 53^ can also not be ruled out for this group and future studies with larger sample sizes are needed to ascertain whether a further refinement of the clinical criteria is needed here.

Aside from the classification of Probable LATE and Possible LATE, the current clinical criteria also allow for a group for whom the clinical symptoms are likely related to co-occurring LATE-NC and AD pathology^1^. In the present study, this LATE-AD group had faster cognitive decline and higher mortality rates compared to “pure” AD, which is in line with previous reports^1, 10, 11, 15, 17, 18^. This accelerated clinical decline is likely driven by the additive burden of both LATE-NC and AD pathology, or the pathological synergy between the two^1, 4, 11, 16, 50, 54^. Aside from the clinical effects of LATE-AD, we also observed that this group shows greater atrophy within the whole brain and an AD-signature cortical thickness mask. While we did not detect accelerated rates of atrophy after our screening visit, this pronounced cortical atrophy in individuals with LATE-AD suggests synergistic pathological effects of LATE-NC and AD pathology on downstream neurodegeneration. Identifying LATE-NC in those who also have AD pathology is currently a major challenge because of the lack of validated molecularly specific biomarkers for TDP-43. What’s more, by allowing the co-occurrence of LATE-NC and AD pathology, one can also not rely on excluding AD biomarkers in this context. Instead, one must rely solely on clinical and radiological features, which is further complicated because these overlap between LATE and AD. The designation of *Possible* LATE-AD, which was not used in this text to avoid confusion with the Possible LATE group, but is the correct classification, therefore, remains very prudent. It is, however, crucial to continue to classify this mixed group as this co-occurring pathology could affect the effectiveness of disease-modifying anti-amyloid medications, which hampers the assessment of potential risks and benefits of these treatments.

Over the years, efforts to identify LATE-NC at autopsy have been published and updated^1, 4, 55^, with the obvious caveat that these cannot be applied during life. Given the current lack of validated biomarkers that can detect LATE-NC *in vivo*, applying clinical criteria predicting LATE-NC is the only way to identify LATE cases during life. The clinical criteria published by Wolk et al, are, however, not the only clinical criteria available. Another set of criteria, published by Corriveau-Lecavalier et al.^50^, aimed to identify a clinical-radiological phenotype that they termed “limbic-predominant amnestic neurodegenerative syndrome” (LANS). Given the similarity in objectives between these two publications, there is a considerable overlap in criteria for LATE^14^ and LANS^50^. The primary conceptual *difference* is that the LATE criteria aim to define a probability that LATE-NC is the etiology of symptoms, while LANS exclusively defines a clinical syndrome. Another dissimilarity between the two is that the LATE criteria rely on the definition of amyloid-β biomarkers, while LANS places more focus on the assessment of neocortical tau using PET, but does not consider amyloid-β biomarkers in the LANS definition. Similarities in specific criteria aside, the conceptual differences between the LATE and LANS definitions could make them suitable for different purposes. LANS could be preferred when only a definition of a clinical entity needs to be defined, and might thereby be more determining in patient management strategies or developing non-pharmacological interventions. However, with recent advances in disease-modifying therapies, it has become apparent that the determining factor for whom to treat will become what pathology is causing the symptoms, rather than targeting the symptomology itself. In this context, and especially when implementing anti-amyloid treatments, the more neurobiologically focused LATE criteria might be more appropriate.

Given the changing therapeutic landscape and the continuing development of biomarkers, clinical criteria for LATE need to be dynamically adapted to accommodate novel findings in the field. Obvious advances in the context of LATE would be molecularly specific markers of TDP-43 and novel genetic markers of LATE-NC. With the addition of each new element to LATE criteria, the accuracy to predict underlying LATE-NC increases. However, it is important to consider that applying these criteria *in vivo* will only ever yield a probability that LATE-NC is the etiology of symptoms, but it is not a definite determination of LATE-NC. A definitive definition of LATE-NC will continue to rely on autopsy data

### Strengths and limitations

A key strength of this study is that we are the first to apply the newly developed clinical criteria for LATE in a large memory clinic cohort, closely resembling the intended real-world clinical use of these criteria. This intended use also guided our decision to classify LATE based on MRI criteria using visual read measures, which are commonly performed in clinical practice and do not require additional MRI processing.

Our study also has several limitations. First, due to the structure of the data source of the Amsterdam Dementia Cohort, which prioritizes comprehensive initial screening over extensive follow-up assessments, we had to operationalize the Wolk et al. 2024^14^ criteria in a way that did not account for the hypothesized indolent course of LATE. However, we were able to demonstrate this progression pattern after classification and show that adapting the criteria to accommodate cross-sectional data results in valid derivative criteria that can be used in memory clinics following the same diagnostic procedures. Second, and similarly, our mean age of 66 (SD:6) years old did not allow us to apply the criteria of “Generally >75 years old”^14^. Instead, we opted to assess age differences between the groups and found that, in our particular sample, older age was not related to a higher chance of LATE. This finding shows that applying these criteria in younger cohorts produces distinct clinical-radiological groups, which could inform both diagnostic procedures and therapeutic strategies for young individuals. Third, our study lacked sufficient tau-PET or FDG-PET data to incorporate these biomarkers into our operationalization of two key supportive features for diagnosing Possible LATE. By primarily assessing CSF p-tau181 rather than tau-PET, which capture distinct biological aspects of tau burden could influence group classification, and tau PET might be preferential due to its specificity to Alzheimer’s disease tau pathology^56^. Previous studies have demonstrated the utility of FDG-PET by assessing inferior temporal vs. medial temporal glucose metabolism ratios^39, 40^, which we could not evaluate in our sample. As a result, our criteria may have been suboptimal to distinguish individuals with LATE-AD from AD, where clinical and radiological features signify the only difference between groups. Fourth, we did not detect any differences in longitudinal atrophy between our diagnostic groups. This is likely due to our limited sample (N=250 across all groups) of individuals with longitudinal MRI, highlighting the need to assess these effects in future large-scale investigations. Fifth, we operationalized and tested clinical criteria for LATE that aim to predict the chance that LATE-NC is the underlying cause for clinical symptoms^14^. However, these criteria have not been thoroughly neuropathological validated, and this needs to be considered when drawing conclusions with regard to clinical *vs* neuropathology associations. Lastly, like many prior studies on LATE, our sample lacks cultural, ethnic, and socioeconomic diversity^57^, limiting the generalizability of our findings. Future research should prioritize recruiting participants from more diverse backgrounds to better reflect the general population.

## Conclusions

LATE presents a significant challenge in neurodegenerative disease research, due to the clinical and radiological similarities to AD, and a definitive diagnosis remains dependent on post-mortem assessment of LATE-NC. As developments in disease-modifying treatments against AD advance, and the possibility that TDP-43 pathology could confound the clinical effects of AD medication, the need to differentiate individuals with LATE and AD pathology becomes imperative. Clinical criteria, such as those discussed here, provide an essential framework for identifying potential LATE cases *in vivo*, yet further advancements in biomarker development are necessary to refine diagnosis and enhance our understanding of the clinical and radiological manifestations of LATE-NC. Exciting advances in biofluid biomarker research, particularly the detection of TDP-43 in plasma extracellular vesicles in frontotemporal dementia (FTD) and amyotrophic lateral sclerosis (ALS) cases, may provide novel avenues for detecting TDP-43 ^58^. These biomarkers could also open up avenues into research on the staging of LATE. Disease staging has been a very active topic of research for AD, and has proven to be a crucial element in determining treatment efficacy in clinical trials. However, these novel biomarkers have not been tested in the context of LATE, and might not be sensitive to all isoforms of TDP-43 pathology. Until valid and reliable biomarkers for detecting LATE-NC during life become available, LATE criteria will continue to rely on clinical and radiological features to obtain a probabilistic estimate of the likelihood of LATE-NC. This study highlights the clinical applicability of these types of criteria, and we hope that further research will continue to advance the field, ultimately leading to accurate and early diagnosis of LATE.

## Abbreviations

AD: Alzheimer’s Disease
ALS: amyotrophic lateral sclerosis
FTLD: fronto-temporal lobar degeneration
MND: motor neuron disease
LATE: Limbic predominant age-related TDP-43 encephalopathy
LANS: limbic-predominant amnestic syndrome
TDP-43: TAR DNA-binding protein 43
APOE: Apolipoprotein E
TMEM106B: Transmembrane protein 106B
MMSE: Mini Mental State Examination
TMT: Trail Making Test
AVLT: Auditory Verbal Learning Test
VAT: Visual Association Test
PET: Positron Emission Tomography
MRI: Magnetic Resonance Imaging
VOSP: Visual Object and Space Perception battery
FAB: Frontal Assessment Battery
ROI: Region of Interest
pg: picogram
MTA: Medial Temporal Atrophy
GCA: Global Cortical Atrophy
PCA: Posterior Cortical Atrophy
ADC: Amsterdam Dementia Cohort

## Acknowledgements and Funding

This work was funded by a Dementia Fellowship grant from Zorg Onderzoek Nederland - Medische Wetenschappen (ZonMW; project number: 10510022110010) and an Alzheimer Nederland Early Career Grant (project number: WE.03-2024-06) to dr. Colin Groot (PI), and a European Research Council (ERC) research grant (project number: 949570) to dr. Rik Ossenkoppele (PI).

Alzheimer Center Amsterdam is supported by Stichting Alzheimer Nederland and Stichting Steun Alzheimercentrum Amsterdam. Rik Ossenkoppele is a recipient of TAP-dementia (TAP-TAU), a ZonMw funded project (#10510032120003) in the context of the Dutch National Dementia Strategy. R.O. has received research funding from European Research Council, ZonMw, NWO, National Institute of Health, Alzheimer Association, Alzheimer Nederland, Stichting Dioraphte, Cure Alzheimer’s fund, Health Holland, ERA PerMed, Alzheimerfonden, Hjarnfonden. I.C. is funded by YOD Molecular.

E.M.C. is supported by Alzheimer Nederland (#WE.03-2024-03).

LEC has acquired research support from GE Healthcare and Springer Healthcare (paid by Eli Lilly), and speakers fee from Life Molecular Imaging, all paid to institution. Dr. Collij is supported by the Alzheimer Association Research Fellowship (#23AARF-1029663) grant.

Research programs of WMvdF have been funded by ZonMW, NWO, EU-JPND, EU-IHI, Alzheimer Nederland, Hersenstichting CardioVascular Onderzoek Nederland, Health∼Holland, Topsector Life Sciences & Health, stichting Dioraphte, Gieskes-Strijbis fonds, stichting Equilibrio, Edwin Bouw fonds, Pasman stichting, stichting Alzheimer & Neuropsychiatrie Foundation, Philips, Biogen MA Inc, Novartis-NL, Life-MI, AVID, Roche BV, Fujifilm, Eisai, Combinostics. WMvdF holds the Pasman chair. WMvdF is recipient of ABOARD, which is a public-private partnership receiving funding from ZonMW (#73305095007) and Health∼Holland, Topsector Life Sciences & Health (PPP-allowance; #LSHM20106). WMvdF is recipient of TAP-dementia (www.tap-dementia.nl), receiving funding from ZonMw (#10510032120003). TAP-dementia receives co-financing from Avid Radiopharmaceuticals and Amprion.

Research of CET is supported by the European Commission (Marie Curie International Training Network, grant agreement No 860197 (MIRIADE) and No 101119596 (TAME), Innovative Medicines Initiatives 3TR (Horizon 2020, grant no 831434) EPND ( IMI 2 Joint Undertaking (JU), grant No. 101034344) and JPND (bPRIDE, CCAD), European Partnership on Metrology, co-financed from the European Union’s Horizon Europe Research and Innovation Programme and by the Participating States ((22HLT07 NEuroBioStand), Horizon Europe (PREDICTFTD, 101156175), CANTATE project funded by the Alzheimer Drug Discovery Foundation, Alzheimer Association, Michael J Fox Foundation, Health Holland, the Dutch Research Council (ZonMW), Alzheimer Drug Discovery Foundation, The Selfridges Group Foundation, Alzheimer Netherlands. CT is recipient of ABOARD, which is a public-private partnership receiving funding from ZonMW (#73305095007) and Health∼Holland, Topsector Life Sciences & Health (PPP-allowance; #LSHM20106). CT is recipient of TAP-dementia, a ZonMw funded project (#10510032120003) in the context of the Dutch National Dementia Strategy.

The funding sources had no role in the design and conduct of the study; in the collection, analysis, interpretation of the data; or in the preparation, review, or approval of the manuscript.

