## Supplement for "Characterizing individuals fulfilling clinical criteria for LATE in a tertiary memory clinic"

| Differences between groups are outlined in Supplemental Table 2 | **Wolk et al. Criteria** | **Operationalization in ADC cohort** |
|---|---|---|
| **I. Core clinical syndrome** | 1. Primary amnestic syndrome with temporo-limbic memory loss **(required)** | Single-domain memory impairment, defined as memory impairment (>2 SDs below mean of controls), with no impairment in any other domain (attention, visuospatial, executive and language). |
| | 2. Other cognitive domains largely spared until much later in the course **(required)** | For individuals with dementia only: Memory domain impairment is more than 1 SD below the patient-specific average across domains, and memory is the most impaired domain. |
| | 3. May have mild semantic memory impairment | Animal fluency (loads on semantic memory) was not included in the language domain when assessing criteria 1 and 2. |
| | 4. Indolent course with predominant amnestic syndrome present for at least 2 years | This is not used as a selection criterion, but assessed in the results. |
| | 5. Age generally > 75 years | This is not used as a selection criterion, but assessed in the results. |
| **II. Required imaging** | Significant hippocampal atrophy (out of proportion to global atrophy) | MTA≥2 (age<75) or MTA≥3 (age≥75) with PCA≤1 and/or GCA≤1,<br>or<br>MTA≥3 (age<75) or MTA ≥4 (age≥75) with PCA≤2 and/or GCA≤2. |
| **III. Required supportive features for Probable LATE** | A negative test of one of the following to rule out amyloid pathology<br>. Amyloid-PET<br>. CSF AB42/40<br>. CSF pTau181/AB42 or t-tau/AB42 | Negative amyloid-β-PET visual read and/or CSFAB42 <813 pg/mL (Innotest, until June 2018) or <1,092 pg/mL (Elecsys, from June 2018 onward). |
| **IV. Required additional measures for Possible LATE if amyloid-positive*** | If amyloid positive based on III, a negative measure of one of the following Tau biomarkers is required:<br>. MTL Tau PET (preferred measure)<br>. CSF pTau181 | Negative on CSF ptau181 or tau-PET visual read |

**Supplemental Table 1. Operationalization of clinical criteria for LATE**
MTA-medial temporal lobe atrophy visual rating score, PCA-posterior cortical atrophy visual rating score, GCA-global cortical atrophy visual rating score, MTL-medial temporal lobe. * $A_{missing}$ individuals were also defined as Possible LATE

| Set of criteria | Criteria | Operationalization in ADC cohort |
|---|---|---|
| **I. Core clinical syndrome** | 1. Progressive amnestic, multi-domain syndrome; memory loss may be particularly severe relative to other cognitive domains **(required)** | Multi-domain impairment including memory and at least one other domain (defined as lower than the mean - 2SD of controls). |
| | 2. Generally more rapid course than typical AD alone **(required)** | Yearly change in MMSE (30 - MMSE score / complaint duration) > 2. |
| **II. Required supportive features for Possible LATE (at least one of the following)** | 1. Severe hippocampal atrophy in the setting of no more than mildly symptomatic dementia | MTA ≥ 3. |
| | 2. MTL/hippocampal atrophy out of proportion to degree of tau pathologic burden measured by tau-PET | Not considered due to limited tau-PET availability |
| | 3. FDG-PET with elevated inferior temporal/MTL ratio | Not considered due to limited FDG-PET availability |

**Supplemental Table 2. Operationalization of clinical criteria for LATE-AD**
MTA-medial temporal lobe atrophy visual rating score, MTL-medial temporal lobe.

| Group: | *Probable LATE vs* | | | *Possible LATE vs* | | *LATE-AD vs* |
|---|---|---|---|---|---|---|
| Reference: | **Possible LATE** | **LATE-AD** | **AD** | **LATE-AD** | **AD** | **AD** |
| **Age** | 0.000 | 0.000 | 1.000 | 0.990 | 0.000 | 0.000 |
| **Sex** | 0.010 | 0.140 | 0.010 | 0.300 | 0.700 | 0.310 |
| **Education** | 0.020 | 0.020 | 0.000 | 0.900 | 0.780 | 0.900 |
| **MMSE** | 0.010 | 0.000 | 0.100 | 0.000 | 0.250 | 0.000 |
| **APOE** | 0.010 | 0.000 | 0.000 | 0.370 | 0.290 | 0.920 |

**Supplemental Table 3. Pairwise differences in demographics between groups**
Differences between groups were assessed using Chi-squared tests (with post hoc Fisher's exact tests), ANOVA (with post hoc Tukey's HSD test), or Kruskal-Wallis tests (with post hoc Dunn's tests), where appropriate. Values displayed are p-values from these tests. Orange cells indicate that the target group is lower than the reference, blue cells indicate the opposite.

*Baseline (LM)*

| Group: | *Probable LATE vs* | | | *Possible LATE vs* | | *LATE-AD vs* |
|---|---|---|---|---|---|---|
| Reference: | **Possible LATE** | **LATE-AD** | **AD** | **LATE-AD** | **AD** | **AD** |
| **MMSE** | 0.65(0.15), 0 | 1.21(0.15), 0 | 0.4(0.13), 0.002 | 0.56(0.12), 0 | -0.25(0.09), 0.007 | -0.8(0.09), 0 |
| **Memory** | 0.42(0.16), 0.008 | 0.32(0.15), 0.039 | -0.41(0.13), 0.002 | -0.1(0.12), 0.419 | -0.83(0.09), 0 | -0.73(0.09), 0 |
| **Executive F.** | 0.69(0.16), 0 | 1.09(0.15), 0 | 0.6(0.13), 0 | 0.4(0.12), 0.001 | -0.09(0.09), 0.307 | -0.5(0.09), 0 |
| **Attention** | 0.45(0.16), 0.004 | 0.98(0.16), 0 | 0.6(0.13), 0 | 0.53(0.12), 0 | 0.15(0.09), 0.11 | -0.38(0.09), 0 |
| **Visuospatial F.** | 0.36(0.19), 0.051 | 1(0.18), 0 | 0.56(0.15), 0 | 0.64(0.15), 0 | 0.19(0.11), 0.091 | -0.44(0.11), 0 |
| **Language** | 0.15(0.17), 0.366 | 0.88(0.16), 0 | 0.26(0.14), 0.056 | 0.73(0.13), 0 | 0.12(0.1), 0.233 | -0.62(0.09), 0 |

*Longitudinal (LMM)*

| Group: | *Probable LATE vs* | | | *Possible LATE vs* | | *LATE-AD vs* |
|---|---|---|---|---|---|---|
| Reference: | **Possible LATE** | **LATE-AD** | **AD** | **LATE-AD** | **AD** | **AD** |
| **MMSE** | 0.1(0.07), 0.16 | 0.24(0.07), 0 | 0.12(0.05), 0.02 | 0.14(0.07), 0.05 | 0.02(0.05), 0.73 | -0.12(0.05), 0.01 |
| **Memory** | 0.07(0.15), 0.64 | 0.34(0.14), 0.01 | 0.24(0.1), 0.01 | 0.27(0.15), 0.07 | 0.17(0.12), 0.16 | -0.1(0.1), 0.3 |
| **Executive F.** | 0.28(0.14), 0.04 | 0.39(0.12), 0 | 0.19(0.09), 0.03 | 0.11(0.14), 0.43 | -0.09(0.11), 0.39 | -0.2(0.09), 0.03 |
| **Attention** | 0.35(0.26), 0.17 | 0.69(0.24), 0 | 0.34(0.17), 0.05 | 0.34(0.25), 0.18 | -0.01(0.19), 0.96 | -0.35(0.17), 0.04 |
| **Visuospatial F.** | 0.37(0.39), 0.33 | 0.39(0.35), 0.26 | 0.43(0.27), 0.1 | 0.02(0.37), 0.96 | 0.06(0.29), 0.84 | 0.04(0.24), 0.88 |
| **Language** | 0.23(0.11), 0.04 | 0.22(0.1), 0.03 | 0.09(0.06), 0.15 | -0.01(0.12), 0.92 | -0.14(0.09), 0.13 | -0.13(0.08), 0.12 |

**Supplemental Table 4. Pairwise differences in cognition between groups**

Baseline effect estimates were obtained using linear regression models, and longitudinal effects (group*time) were obtained using linear mixed-effects analyses. All models were corrected for age, sex, and education. Blue shaded cells indicate a significant positive effect (higher scores in target group), orange cells indicate a significant negative effect. LM=linear model, LMM=linear mixed effects model.

| Group: | *Probable LATE vs* | | | *Possible LATE vs* | | *LATE-AD vs* |
|---|---|---|---|---|---|---|
| Reference: | **Possible LATE** | **LATE-AD** | **AD** | **LATE-AD** | **AD** | **AD** |
| **Hazard ratio** | 1.68[1.12-2.54], 0.013 | 1.79[1.2-2.68], 0.004 | 1.44[1.01-2.05], 0.044 | 1.07[0.8-1.42], 0.665 | 0.85[0.68-1.07], 0.174 | 0.8[0.65-0.99], 0.039 |

**Supplemental Table 5. Differences in mortality rates between groups**

Hazard ratios were obtained using Cox proportional hazard models, which were corrected for age, sex, education, and syndrome diagnosis. Blue shaded cells indicate a significant positive effect (lower mortality risk in target group), orange cells indicate a significant negative effect.

*Baseline (LM)*

| Group: | *Probable LATE vs* | | | *Possible LATE vs* | | *LATE-AD vs* |
|---|---|---|---|---|---|---|
| Reference: | **Possible LATE** | **LATE-AD** | **AD** | **LATE-AD** | **AD** | **AD** |
| **Whole-brain** | -0.06(0.2), 0.784 | -0.7(0.2), 0 | -0.06(0.16), 0.699 | -0.65(0.17), 0 | -0.01(0.13), 0.956 | 0.64(0.12), 0 |
| **AD signature** | -0.16(0.2), 0.442 | -0.9(0.19), 0 | -0.16(0.16), 0.31 | -0.74(0.17), 0 | -0.01(0.13), 0.945 | 0.73(0.11), 0 |
| **Hippocampus** | 0.47(0.18), 0.011 | 0.46(0.18), 0.01 | 1.04(0.15), 0 | -0.01(0.15), 0.953 | 0.57(0.12), 0 | 0.58(0.1), 0 |
| **Amygdala** | 0.12(0.19), 0.524 | -0.07(0.18), 0.69 | 0.55(0.15), 0 | -0.19(0.15), 0.213 | 0.43(0.12), 0 | 0.62(0.11), 0 |
| **Entorhinal** | 0.33(0.19), 0.079 | 0.3(0.18), 0.101 | 0.33(0.15), 0.028 | -0.04(0.15), 0.821 | 0(0.12), 0.992 | 0.04(0.11), 0.734 |
| **ITH ratio** | -0.18(0.2), 0.366 | -0.47(0.19), 0.015 | -0.59(0.16), 0 | -0.29(0.17), 0.081 | -0.4(0.13), 0.002 | -0.11(0.11), 0.321 |

*Longitudinal (LMM)*

| Group: | *Probable LATE vs* | | | *Possible LATE vs* | | *LATE-AD vs* |
|---|---|---|---|---|---|---|
| Reference: | **Possible LATE** | **LATE-AD** | **AD** | **LATE-AD** | **AD** | **AD** |
| **Whole-brain** | 0.15(0.19), 0.438 | -0.24(0.19), 0.214 | -0.11(0.07), 0.134 | -0.39(0.26), 0.125 | -0.26(0.18), 0.151 | 0.13(0.18), 0.468 |
| **AD signature** | 1.12(0.21), 0.545 | 0.76(0.14), 0.137 | 0.9(0.07), 0.166 | 0.68(0.16), 0.109 | 0.81(0.14), 0.215 | 1.19(0.2), 0.31 |
| **Hippocampus** | 0.07(0.11), 0.515 | 0.13(0.11), 0.241 | 0.02(0.05), 0.737 | 0.05(0.14), 0.701 | -0.06(0.1), 0.578 | -0.11(0.1), 0.26 |
| **Amygdala** | 0.14(0.12), 0.243 | 0.02(0.12), 0.835 | -0.02(0.05), 0.654 | -0.12(0.15), 0.438 | -0.17(0.11), 0.134 | -0.05(0.11), 0.647 |
| **Entorhinal** | -0.15(0.11), 0.168 | -0.09(0.11), 0.39 | -0.07(0.04), 0.104 | 0.06(0.14), 0.691 | 0.08(0.1), 0.41 | 0.03(0.1), 0.79 |
| **ITH ratio** | -0.15(0.14), 0.281 | -0.22(0.14), 0.107 | -0.08(0.06), 0.141 | -0.07(0.18), 0.693 | 0.07(0.13), 0.594 | 0.14(0.13), 0.27 |

**Supplemental Table 6. Baseline and longitudinal differences in atrophy between diagnostic groups**
Effect estimates obtained using linear regression models (baseline effects) and linear mixed-effects analyses, which were corrected for age, and sex. Values displayed are β(SE), p for the group (baseline) and group*time interaction (longitudinal) effects. Blue shaded cells indicate a significant positive effect (higher baseline values, or slower longitudinal decline), orange cells indicate a significant negative

| | | | | |
|---|---|---|---|---|
| Baseline (LM) | | | | |
| Group: | *Probable LATE* | | | |
| Reference: | **Possible LATE A+T-** | **Possible LATE Amissing** | **LATE-AD** | **AD** |
| **MMSE** | -0,5(0,19), 0,007 | -0,68(0,17), 0 | -1,09(0,15), 0 | -0,34(0,13), 0,008 |
| **Memory** | -0,24(0,18), 0,182 | -0,55(0,17), 0,001 | -0,37(0,15), 0,013 | 0,34(0,12), 0,006 |
| **Executive F.** | -0,6(0,19), 0,001 | -0,8(0,18), 0 | -1,1(0,16), 0 | -0,59(0,13), 0 |
| **Attention** | -0,38(0,19), 0,051 | -0,67(0,18), 0 | -0,93(0,16), 0 | -0,63(0,13), 0 |
| **Visuospatial F.** | -0,28(0,21), 0,18 | -0,48(0,22), 0,028 | -0,84(0,18), 0 | -0,5(0,14), 0,001 |
| **Language** | 0,01(0,19), 0,975 | -0,06(0,18), 0,719 | -0,72(0,16), 0 | -0,16(0,13), 0,228 |
| *Longitudinal (LMM)* | | | | |
| Group: | *Probable LATE* | | | |
| Reference: | **Possible LATE A+T-** | **Possible LATE Amissing** | **LATE-AD** | **AD** |
| **MMSE** | -0,12(0,06), 0,032 | -0,09(0,06), 0,121 | -0,17(0,04), 0 | -0,07(0,03), 0,017 |
| **Memory** | -0,02(0,07), 0,71 | -0,1(0,08), 0,183 | -0,18(0,05), 0 | -0,1(0,03), 0,001 |
| **Executive F.** | -0,19(0,06), 0,002 | -0,03(0,08), 0,681 | -0,16(0,04), 0 | -0,06(0,03), 0,02 |
| **Attention** | -0,07(0,07), 0,284 | -0,09(0,08), 0,268 | -0,19(0,05), 0 | -0,05(0,03), 0,101 |
| **Visuospatial F.** | -0,15(0,09), 0,078 | 0(0,18), 0,985 | -0,14(0,07), 0,051 | -0,1(0,05), 0,049 |
| **Language** | -0,01(0,07), 0,944 | -0,32(0,09), 0 | -0,12(0,06), 0,03 | -0,05(0,03), 0,147 |

| | | | |
|---|---|---|---|
| *Baseline (LM)* | *Possible LATE A+T-* | | |
| Group: | **Possible LATE Amissing** | **LATE-AD** | **AD** |
| Reference: | -0,17(0,18), 0,339 | -0,59(0,16), 0 | 0,16(0,14), 0,244 |
| **MMSE** | -0,31(0,18), 0,081 | -0,13(0,16), 0,418 | 0,58(0,14), 0 |
| **Memory** | -0,21(0,18), 0,264 | -0,51(0,16), 0,002 | 0,01(0,14), 0,955 |
| **Executive F.** | -0,29(0,19), 0,123 | -0,56(0,17), 0,001 | -0,25(0,14), 0,085 |
| **Attention** | -0,2(0,23), 0,365 | -0,56(0,18), 0,002 | -0,22(0,15), 0,145 |
| **Visuospatial F.** | -0,07(0,19), 0,706 | -0,73(0,17), 0 | -0,16(0,14), 0,247 |
| **Language** | | | |
| *Longitudinal (LMM)* | *Possible LATE A+T-* | | |
| Group: | **Possible LATE Amissing** | **LATE-AD** | **AD** |
| Reference: | 0,03(0,07), 0,66 | -0,04(0,06), 0,453 | 0,06(0,05), 0,27 |
| **MMSE** | -0,08(0,09), 0,389 | -0,15(0,07), 0,03 | -0,07(0,06), 0,215 |
| **Memory** | 0,16(0,1), 0,098 | 0,03(0,07), 0,605 | 0,13(0,06), 0,019 |
| **Executive F.** | -0,02(0,1), 0,868 | -0,11(0,07), 0,134 | 0,02(0,06), 0,73 |
| **Attention** | 0,16(0,18), 0,391 | 0,01(0,09), 0,885 | 0,06(0,07), 0,44 |
| **Visuospatial F.** | -0,32(0,1), 0,002 | -0,12(0,08), 0,145 | -0,05(0,07), 0,498 |

| *Baseline (LM)* | *Possible LATE Amissing vs* | | *LATE-AD vs* |
|---|---|---|---|
| Group: | **LATE-AD** | **AD** | **AD** |
| Reference: | -0,41(0,15), 0,005 | 0,34(0,12), 0,006 | 0,75(0,09), 0 |
| **MMSE** | 0,18(0,14), 0,205 | 0,89(0,12), 0 | 0,71(0,09), 0 |
| **Memory** | -0,3(0,15), 0,044 | 0,21(0,12), 0,085 | 0,51(0,09), 0 |
| **Executive F.** | -0,26(0,15), 0,086 | 0,04(0,13), 0,731 | 0,31(0,09), 0,001 |
| **Attention** | -0,35(0,2), 0,073 | -0,02(0,17), 0,911 | 0,33(0,11), 0,002 |
| **Visuospatial F.** | -0,66(0,15), 0 | -0,09(0,13), 0,457 | 0,56(0,09), 0 |
| **Language** | | | |
| *Longitudinal (LM)* | *Possible LATE Amissing vs* | | *LATE-AD vs* |
| Group: | **LATE-AD** | **AD** | **AD** |
| Reference: | -0,08(0,06), 0,204 | 0,02(0,05), 0,643 | 0,1(0,03), 0,001 |
| **MMSE** | -0,07(0,08), 0,376 | 0,01(0,07), 0,919 | 0,08(0,04), 0,045 |
| **Memory** | -0,12(0,09), 0,147 | -0,03(0,08), 0,731 | 0,1(0,04), 0,007 |
| **Executive F.** | -0,1(0,09), 0,266 | 0,04(0,08), 0,619 | 0,13(0,04), 0,002 |
| **Attention** | -0,15(0,18), 0,414 | -0,1(0,17), 0,551 | 0,04(0,05), 0,421 |
| **Visuospatial F.** | 0,2(0,09), 0,031 | 0,27(0,08), 0,001 | 0,07(0,05), 0,116 |
| **Language** | | | |

**Supplemental Table 7. Pairwise differences in cognition between groups, Possible LATE A+T- and Possible LATE Amissing separated**
Effect estimates obtained using linear regression models (baseline effects) and linear mixed-effects analyses, which were corrected for age, sex, and education. Values displayed are β(SE), p for the group (baseline) and group*time interaction (longitudinal) effects. Blue shaded cells indicate a significant positive effect (higher baseline values, or slower longitudinal decline), orange cells indicate a significant negative

*Baseline (LM)*

| | | | | |
|---|---|---|---|---|
| Group: | <u>*Probable LATE*</u> | | | |
| Reference: | **Probable LATE T+** | **Possible LATE** | **LATE-AD** | **AD** |
| **MMSE** | -0.21(0.26), 0.419 | -0.69(0.18), 0 | -1.18(0.18), 0 | -0.43(0.16), 0.008 |
| **Memory** | -0,19(0,26), 0,477 | -0,79(0,19), 0 | -1,18(0,18), 0 | -0,67(0,16), 0 |
| **Executive F.** | -0,06(0,27), 0,835 | -0,56(0,19), 0,003 | -0,95(0,19), 0 | -0,65(0,17), 0 |
| **Attention** | -0,19(0,29), 0,518 | -0,47(0,21), 0,028 | -0,94(0,21), 0 | -0,6(0,18), 0,001 |
| **Visuospatial F.** | 0,08(0,27), 0,778 | -0,01(0,19), 0,94 | -0,7(0,18), 0 | -0,14(0,16), 0,405 |
| **Language** | -0,07(0,25), 0,774 | -0,44(0,18), 0,013 | -0,39(0,18), 0,025 | 0,32(0,16), 0,045 |

*Longitudinal (LM)*

| | | | | |
|---|---|---|---|---|
| Group: | <u>*Probable LATE*</u> | | | |
| Reference: | **Probable LATE T+** | **Possible LATE** | **LATE-AD** | **AD** |
| **MMSE** | -0.07(0.06), 0.27 | -0.12(0.05), 0.011 | -0.18(0.04), 0 | -0.08(0.03), 0.009 |
| **Memory** | 0,02(0,07), 0,824 | -0,05(0,06), 0,419 | -0,17(0,05), 0,001 | -0,09(0,03), 0,007 |
| **Executive F.** | -0,04(0,06), 0,556 | -0,14(0,05), 0,007 | -0,17(0,05), 0 | -0,07(0,03), 0,021 |
| **Attention** | 0,01(0,07), 0,924 | -0,07(0,06), 0,215 | -0,18(0,06), 0,001 | -0,05(0,04), 0,164 |
| **Visuospatial F.** | -0,23(0,15), 0,134 | -0,15(0,08), 0,08 | -0,16(0,07), 0,028 | -0,12(0,05), 0,023 |
| **Language** | -0,07(0,08), 0,402 | -0,14(0,06), 0,025 | -0,14(0,06), 0,021 | -0,07(0,04), 0,095 |

*Baseline (LM)*

| Group: | *Probable LATE T+ vs* | | |
|---|---|---|---|
| Reference: | **Possible LATE** | **LATE-AD** | **AD** |
| **MMSE** | -0.48(0.22), 0.032 | -0.97(0.22), 0 | -0.22(0.21), 0.285 |
| **Memory** | -0,6(0,23), 0,009 | -0,99(0,23), 0 | -0,48(0,21), 0,023 |
| **Executive F.** | -0,51(0,23), 0,031 | -0,9(0,23), 0 | -0,59(0,22), 0,006 |
| **Attention** | -0,28(0,26), 0,282 | -0,75(0,25), 0,003 | -0,41(0,23), 0,077 |
| **Visuospatial F.** | -0,09(0,23), 0,7 | -0,77(0,23), 0,001 | -0,21(0,21), 0,322 |
| **Language** | -0,37(0,22), 0,093 | -0,32(0,22), 0,142 | 0,39(0,2), 0,056 |

*Longitudinal (LMM)*

| Group: | *Probable LATE vs* | | |
|---|---|---|---|
| Reference: | **Possible LATE** | **LATE-AD** | **AD** |
| **MMSE** | -0.05(0.07), 0.444 | -0.11(0.06), 0.079 | -0.01(0.06), 0.837 |
| **Memory** | -0,06(0,08), 0,42 | -0,19(0,07), 0,009 | -0,11(0,06), 0,076 |
| **Executive F.** | -0,11(0,07), 0,119 | -0,13(0,06), 0,042 | -0,03(0,05), 0,541 |
| **Attention** | -0,08(0,08), 0,312 | -0,19(0,08), 0,013 | -0,06(0,06), 0,372 |
| **Visuospatial F.** | 0,08(0,16), 0,617 | 0,06(0,15), 0,678 | 0,11(0,14), 0,453 |
| **Language** | -0,08(0,09), 0,378 | -0,07(0,08), 0,402 | 0(0,07), 0,986 |

*Baseline (LM)*

| Group: | *Possible LATE vs* | | *LATE-AD vs* |
|---|---|---|---|
| Reference: | **LATE-AD** | **AD** | **AD** |
| **MMSE** | -0.49(0.12), 0 | 0.26(0.09), 0.005 | 0.75(0.09), 0 |
| **Memory** | -0,39(0,13), 0,002 | 0,12(0,09), 0,2 | 0,51(0,09), 0 |
| **Executive F.** | -0,39(0,13), 0,003 | -0,08(0,1), 0,389 | 0,31(0,09), 0,001 |
| **Attention** | -0,47(0,15), 0,002 | -0,14(0,12), 0,24 | 0,33(0,11), 0,002 |
| **Visuospatial F.** | -0,68(0,13), 0 | -0,12(0,1), 0,2 | 0,56(0,09), 0 |
| **Language** | 0,05(0,12), 0,693 | 0,76(0,09), 0 | 0,71(0,09), 0 |

*Longitudinal (LMM)*

| Group: | *Possible LATE vs* | | *LATE-AD vs* |
|---|---|---|---|
| Reference: | **LATE-AD** | **AD** | **AD** |
| **MMSE** | -0.06(0.05), 0.195 | 0.04(0.04), 0.28 | 0.1(0.03), 0.001 |
| **Memory** | -0,13(0,06), 0,034 | -0,05(0,05), 0,3 | 0,08(0,04), 0,045 |
| **Executive F.** | -0,02(0,06), 0,708 | 0,08(0,05), 0,096 | 0,1(0,04), 0,007 |
| **Attention** | -0,11(0,06), 0,084 | 0,02(0,05), 0,631 | 0,13(0,04), 0,002 |
| **Visuospatial F.** | -0,02(0,09), 0,858 | 0,03(0,07), 0,674 | 0,04(0,05), 0,421 |
| **Language** | 0,01(0,07), 0,921 | 0,08(0,05), 0,132 | 0,07(0,05), 0,118 |

**Supplemental Table 8. Pairwise differences in cognition between groups, Probable LATE and Probable LATE T+ separated**
Effect estimates obtained using linear regression models (baseline effects) and linear mixed-effects analyses, which were corrected for age, sex, and education. Values displayed are β(SE), p for the group (baseline) and group*time interaction (longitudinal) effects. Blue shaded cells indicate a significant positive effect (higher baseline values, or slower longitudinal decline), orange cells indicate a significant negative

| Reference: | *Probable LATE* | | | | *Possible LATE A+T-* | | |
|---|---|---|---|---|---|---|---|
| Group: | **Possible LATE A+T-** | **Possible LATE Amissing** | **LATE-AD** | **AD** | **Possible LATE Amissing** | **LATE-AD** | **AD** |
| **Hazard Ratio** | 1.56[0.94-2.58], 0.086 | 1.76[1.13-2.73], 0.012 | 1.8[1.2-2.68], 0.004 | 1.44[1.01-2.05], 0.044 | 1.13[0.72-1.78], 0.595 | 1.15[0.76-1.75], 0.5 | 0.92[0.64-1.34], 0.67 |

**Supplemental Table 9. HR differences between diagnostics groups, Possible LATE A+T- and Possible LATE Amissing separated**

Hazard ratios obtained using Cox proportional hazard models, which were corrected for age, sex, education and syndrome diagnosis. Blue shaded cells indicate a significant positive effect (lower mortality risk in target group), orange cells indicate a significant negative effect.

| Reference: | *Probable LATE* | | | | *Probable LATE T+ vs* | | | *Possib* |
|---|---|---|---|---|---|---|---|---|
| Group: | **Probable LATE T+** | **Possible LATE** | **LATE-AD** | **AD** | **Possible LATE** | **LATE-AD** | **AD** | **LATE-** |
| **HR** | 0.35[0.15-0.82], 0.015 | 1.2[0.77-1.88], 0.422 | 1.28[0.82-1.99], 0.269 | 1.03[0.69-1.53], 0.895 | 3.4[1.57-7.37], 0.002 | 3.63[1.68-7.82], 0.001 | 2.91[1.38-6.12], 0.005 | 1.07[0 |

**Supplemental Table 10. HR differences between diagnostics groups, Probable LATE and Probable LATE T+ separated**

Hazard ratios obtained using Cox proportional hazard models, which were corrected for age, sex, education and syndrome diagnosis. Blue shaded cells indicate a significant positive effect (lower mortality risk in target group), orange cells indicate a significant negative effect.

*Baseline (LM)*

| Reference: | *Probable LATE* | | | | *Possible LATE A+T-* | | | *Possible LATE Amissing vs* | | *LATE-AD vs* |
|---|---|---|---|---|---|---|---|---|---|---|
| Group: | **Possible LATE A+T-** | **Possible LATE Amissing** | **LATE-AD** | **AD** | **Possible LATE Amissing** | **LATE-AD** | **AD** | **LATE-AD** | **AD** | **AD** |
| **Whole-brain** | 0,24(0,46), 0,604 | -0,12(0,95), 0,897 | -0,45(0,42), 0,291 | -0,11(0,07), 0,134 | -0,36(0,96), 0,709 | -0,68(0,45), 0,127 | 0,08(0,34), 0,823 | -0,32(0,94), 0,731 | 0,44(0,9), 0,629 | 0,76(0,3), 0,011 |
| **AD signature** | 0,27(0,45), 0,553 | -0,24(0,93), 0,792 | -0,59(0,42), 0,16 | -0,05(0,07), 0,526 | -0,51(0,94), 0,587 | -0,85(0,44), 0,053 | 0,33(0,34), 0,331 | -0,34(0,92), 0,71 | 0,84(0,88), 0,341 | 1,18(0,29), 0 |
| **Hippocampus** | 0,61(0,39), 0,121 | 1,46(0,29), 0 | 0,14(0,16), 0,377 | 0,06(0,86), 0,944 | 0,67(0,31), 0,034 | 0,08(0,12), 0,478 | -0,03(0,33), 0,93 | -0,11(0,34), 0,746 | 0,04(0,15), 0,76 | 0,04(0,33), 0,891 |
| **Amygdala** | 0,44(0,43), 0,306 | 0,86(0,32), 0,008 | -0,11(0,1), 0,26 | 0,45(0,94), 0,634 | 0,55(0,34), 0,113 | 0,12(0,13), 0,339 | 0,33(0,35), 0,348 | 0,21(0,37), 0,57 | -0,1(0,16), 0,534 | - 0,36(0,35), 0,309 |
| **Entorhinal** | -0,17(0,44), 0,71 | 0,2(0,33), 0,544 | -0,05(0,11), 0,647 | -0,5(0,98), 0,609 | 0,24(0,35), 0,499 | -0,15(0,11), 0,178 | -0,13(0,34), 0,696 | 0,02(0,36), 0,966 | 0,06(0,14), 0,686 | 0,07(0,34), 0,84 |
| **ITH ratio** | -1(0,4), 0,013 | -1,29(0,3), 0 | 0,03(0,1), 0,79 | -0,32(0,88), 0,719 | -0,96(0,32), 0,003 | -0,18(0,15), 0,226 | 0,11(0,44), 0,8 | 0,29(0,45), 0,526 | - 0,05(0,19), 0,803 | - 0,19(0,43), 0,656 |

*Longitudinal (LMM)*

| Reference: | *Probable LATE* | | | | *Possible LATE A+T-* | | | *Possible LATE Amissing vs* | | *LATE-AD vs* |
|---|---|---|---|---|---|---|---|---|---|---|
| Group: | **Possible LATE A+T-** | **Possible LATE Amissing** | **LATE-AD** | **AD** | **Possible LATE Amissing** | **LATE-AD** | **AD** | **LATE-AD** | **AD** | **AD** |
| **Whole-brain** | 0,24(0,46), 0,604 | -0,12(0,95), 0,897 | -0,45(0,42), 0,291 | 0,31(0,31), 0,317 | -0,36(0,96), 0,709 | -0,68(0,45), 0,127 | 0,08(0,34), 0,823 | -0,32(0,94), 0,731 | 0,44(0,9), 0,629 | 0,76(0,3), 0,011 |
| **AD signature** | 0,27(0,45), 0,553 | -0,24(0,93), 0,792 | -0,59(0,42), 0,16 | 0,59(0,31), 0,055 | -0,51(0,94), 0,587 | -0,85(0,44), 0,053 | 0,33(0,34), 0,331 | -0,34(0,92), 0,71 | 0,84(0,88), 0,341 | 1,18(0,29), 0 |
| **Hippocampus** | 0,61(0,39), 0,121 | 1,46(0,29), 0 | 0,14(0,16), 0,377 | 1,18(0,29), 0 | 0,67(0,31), 0,034 | 0,08(0,12), 0,478 | -0,03(0,33), 0,93 | -0,11(0,34), 0,746 | 0,04(0,15), 0,76 | 0,04(0,33), 0,891 |
| **Amygdala** | 0,44(0,43), 0,306 | 0,86(0,32), 0,008 | -0,11(0,1), 0,26 | 0,85(0,28), 0,002 | 0,55(0,34), 0,113 | 0,12(0,13), 0,339 | 0,33(0,35), 0,348 | 0,21(0,37), 0,57 | -0,1(0,16), 0,534 | - 0,36(0,35), |

| | | | | | | | | | | |
|---|---|---|---|---|---|---|---|---|---|---|
| | | | | | | | | | | 0,309 |
| **Entorhinal** | -0,17(0,44), 0,71 | 0,2(0,33), 0,544 | -0,05(0,11), 0,647 | 0,42(0,3), 0,168 | 0,24(0,35), 0,499 | -0,15(0,11), 0,178 | -0,13(0,34), 0,696 | 0,02(0,36), 0,966 | 0,06(0,14), 0,686 | 0,07(0,34), 0,84 |
| **ITH ratio** | -1(0,4), 0,013 | -1,29(0,3), 0 | 0,03(0,1), 0,79 | 0,37(0,31), 0,242 | -0,96(0,32), 0,003 | -0,18(0,15), 0,226 | 0,11(0,44), 0,8 | 0,29(0,45), 0,526 | -0,05(0,19), 0,803 | -0,19(0,43), 0,656 |

**Supplemental Table 11. Pairwise differences in atrophy between groups, Possible LATE A+T- and Possible LATE Amissing separated**

Effect estimates obtained using linear regression models (baseline effects) and linear mixed-effects analyses, which were corrected for age, and sex. Values displayed are β(SE), p for the group (baseline) and group*time interaction (longitudinal) effects. Blue shaded cells indicate a significant positive effect (higher baseline values, or slower longitudinal decline), orange cells indicate a significant negative

**A**

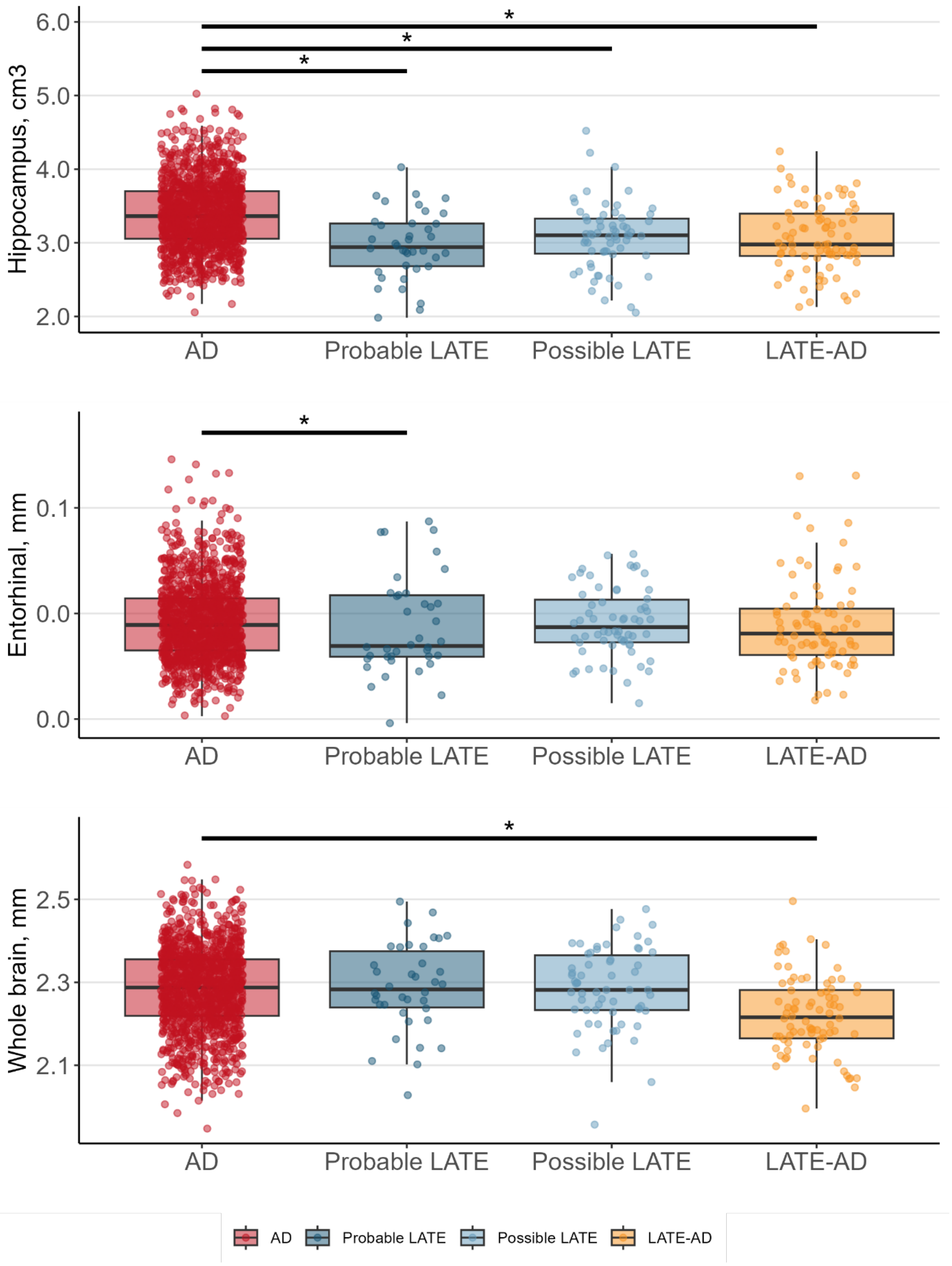

Hippocampus, cm3
6.0
5.0
4.0
3.0
2.0
*
*
*
AD
Probable LATE
Possible LATE
LATE-AD
Entorhinal, mm
0.1
0.0
0.0
*
AD
Probable LATE
Possible LATE
LATE-AD
Whole brain, mm
2.5
2.3
2.1
*
AD
Probable LATE
Possible LATE
LATE-AD
AD
Probable LATE
Possible LATE
LATE-AD

**B**

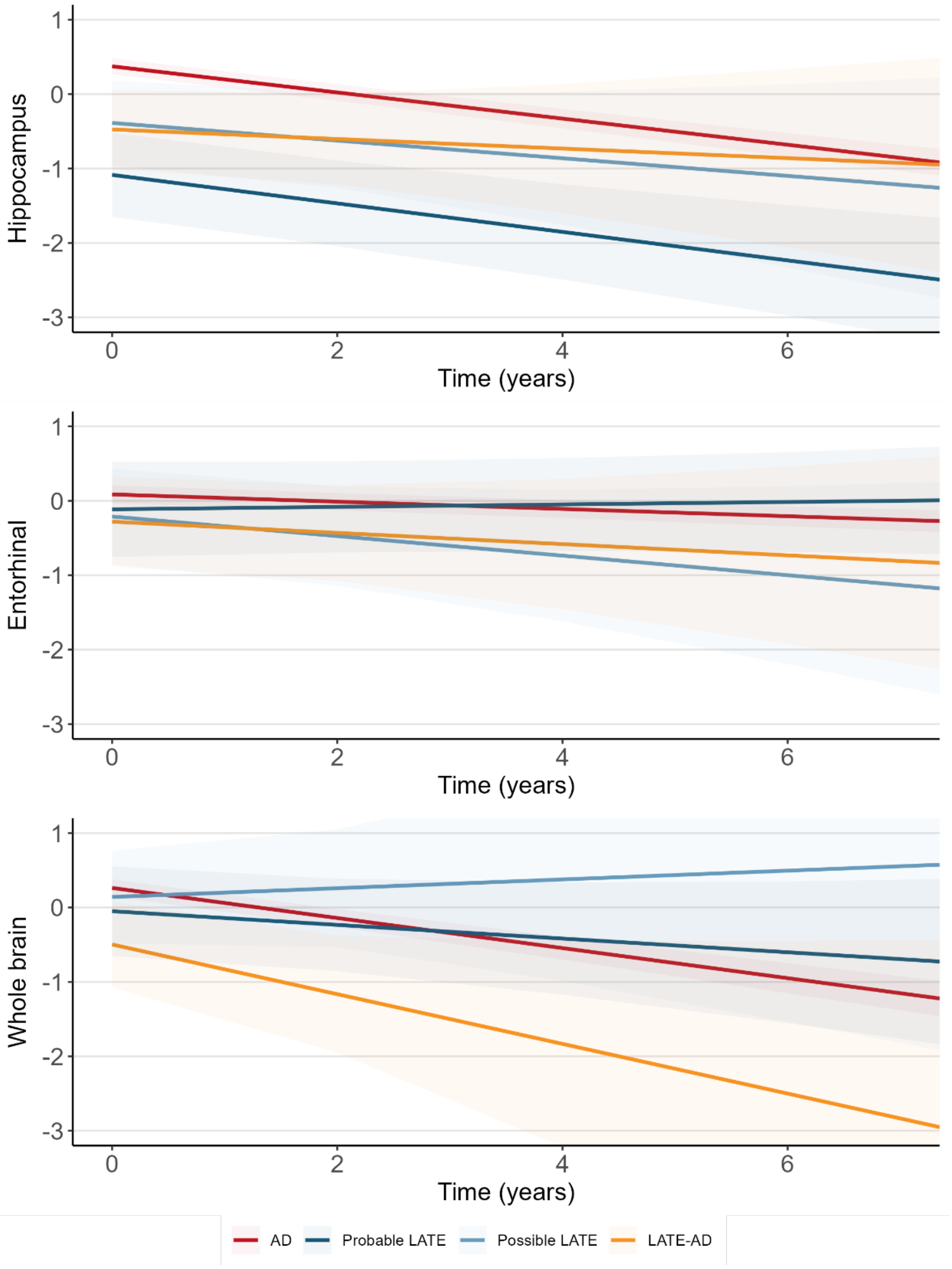


**Supplemental Figure 1. Baseline (A) and longitudinal (B) brain atrophy across diagnostic groups**
(A) Bar graph showing group differences in baseline atrophy. Differences between AD and the LATE groups obtained using general linear models are indicated by the significance bars.(B) Line graphs indicate differences in longitudinal atrophy between groups. Differences between AD and the LATE groups obtained using linear mixed effects analyses (indicated by the slope) are highlighted with an * (none were significant). All pairwise differences between groups are outlined in Supplemental Table 6 ITH=inferior temporal thickness/hippocampal volume, higher values indicate more limbic-predominant atrophy.
